# Development and validation of a parent proxy bronchiectasis child quality of life instrument: The BC-QoL

**DOI:** 10.64898/2025.12.15.25341944

**Authors:** Jack M. Roberts, Peter A. Newcombe, Vikas Goyal, Sanjeewa Kularatna, Steven M. McPhail, Anne B. Chang, Julie M. Marchant

## Abstract

**Background:** Quality of life (QoL), the highest prioritised outcome by children with bronchiectasis and their parents, is a patient-reported outcome measure increasingly considered essential when evaluating health and interventions. Despite this, there are no validated instruments that specifically measure QoL related to bronchiectasis in children. We aimed to develop and validate a new bronchiectasis child-specific parent-proxy QoL instrument (BC-QoL).

**Methods:** We developed a draft 44-item BC-QoL and subsequently conducted a prospective cohort study where 142 parents completed draft BC-QoL and six other measures: two validated cough scores, parent-proxy children’s acute cough-specific QoL, depression, anxiety and stress 21-item scale, RAND-36 and paediatric QoL (PedsQL^TM^4.0) to assess the convergent and discriminant validity of the BC-QoL. The questionnaires were completed over three weeks at different phases of their child’s illness (stable state, exacerbation and/or recovery). Responses were analysed using psychometric and clinical impact techniques to reduce items and determine the instrument’s reliability and validity. Minimally important difference (MID) was also calculated.

**Results:** The final 23-item BC-QoL instrument with its three domains (emotional, physical, social well-being) demonstrated high split half reliability (0.95), Cronbach’s alpha (0.97), repeatability (intraclass coefficient=0.74, 95%CI 0.62-0.82), validity and responsiveness. BC-QoL’s domains significantly correlated with that of PedsQL^TM^4.0 (Spearman correlations: emotional=0.43, social=0.41, physical=0.47). BC-QoL’s MID ranged from 0.74 to 1.32.

**Conclusion:** BC-QoL, the first bronchiectasis child-specific QoL instrument, has evidence demonstrating its validity and reliability, and can be used to evaluate the impact and effectiveness of interventions and better understand the disease burden in children with bronchiectasis.

## Introduction

Worldwide, childhood bronchiectasis has been increasingly recognised as a chronic illness of increasing prevalence with substantial morbidity,[1,2] especially regarding its substantial negative impact on the quality of life (QoL) of children and their carers.[3,4] The costs associated with bronchiectasis to healthcare systems is also substantial, with annual health related costs in the USA estimated at US$14.68 billion in adults.[5] With renewed interest in bronchiectasis, clinical trials of novel interventions have been undertaken to attempt to ameliorate the burden of the disease and improve short- and long-term outcomes, primarily by reducing the frequency, duration and severity of exacerbations.[6–8] To advance the quality of clinical trials in ways that are relevant and meaningful to patients, patient-reported outcome measures (PROMs) are required, and these, especially those measuring health-related QoL (HR-QoL), are increasingly considered essential for evaluating both health and interventions.[9] In childhood bronchiectasis, HR-QoL was the highest rated outcome by parents in an international consensus study and forms a core outcome in clinical studies.[9] However, no disease-specific instrument for measuring HR-QoL in children with bronchiectasis currently exists.[3]

Disease-specific and age-appropriate HR-QoL instruments are typically more sensitive than generic ones, especially in chronic conditions,[10] and are increasingly considered necessary for a holistic representation of HR-QoL outcomes in conjunction with generic measures.[11] Consequently, many disease and condition specific QoL instruments for children are currently in use (e.g. in diabetes,[12] cerebral palsy,[13] and cardiac conditions[14]). Several paediatric pulmonology-specific HR-QoL instruments also exist, including the paediatric asthma caregiver’s QoL questionnaire (PACQLQ),[15] parent-proxy child chronic cough QoL (PC-QoL),[16–18] and parent-proxy children’s acute cough-specific QoL (PAC-QoL).[19] Whilst, chronic cough is the cardinal symptom of bronchiectasis[1,3] leading to use of the cough specific PC-QoL in paediatric bronchiectasis studies,[6] the many other symptoms and impact of bronchiectasis beyond cough remain under-accounted for. Consequently, the QoL-Bronchiectasis (QoL-B)[20] and Bronchiectasis Health Questionnaire (BHQ)[21] have been developed for adults with bronchiectasis. However, neither is optimal or appropriate for use with young children, primarily because they both have domains specifically concerning work and household activities, but also because neither questionnaire was developed with children in mind.

In the absence of a child-specific bronchiectasis HR-QoL and the clinician and consumer identified need for one,[4,9] we aimed to co-develop, validate, and estimate the minimally important difference (MID) of a bronchiectasis child-specific parent-proxy QoL questionnaire (BC-QoL).

## Methods

We used a step-wise approach in the development and validation of the BC-QoL, as previously done.[22–24] Figure 1 depicts the overall methodology with each step further detailed in e-appendix-1. The study was approved by the research ethics committees of Children’s Health Queensland (HREC/20/QCHQ/64160) and the Queensland University of Technology (2000000433), Australia. Written informed consent was obtained from parents and assent from children ≥12 years.

**Figure 1:**
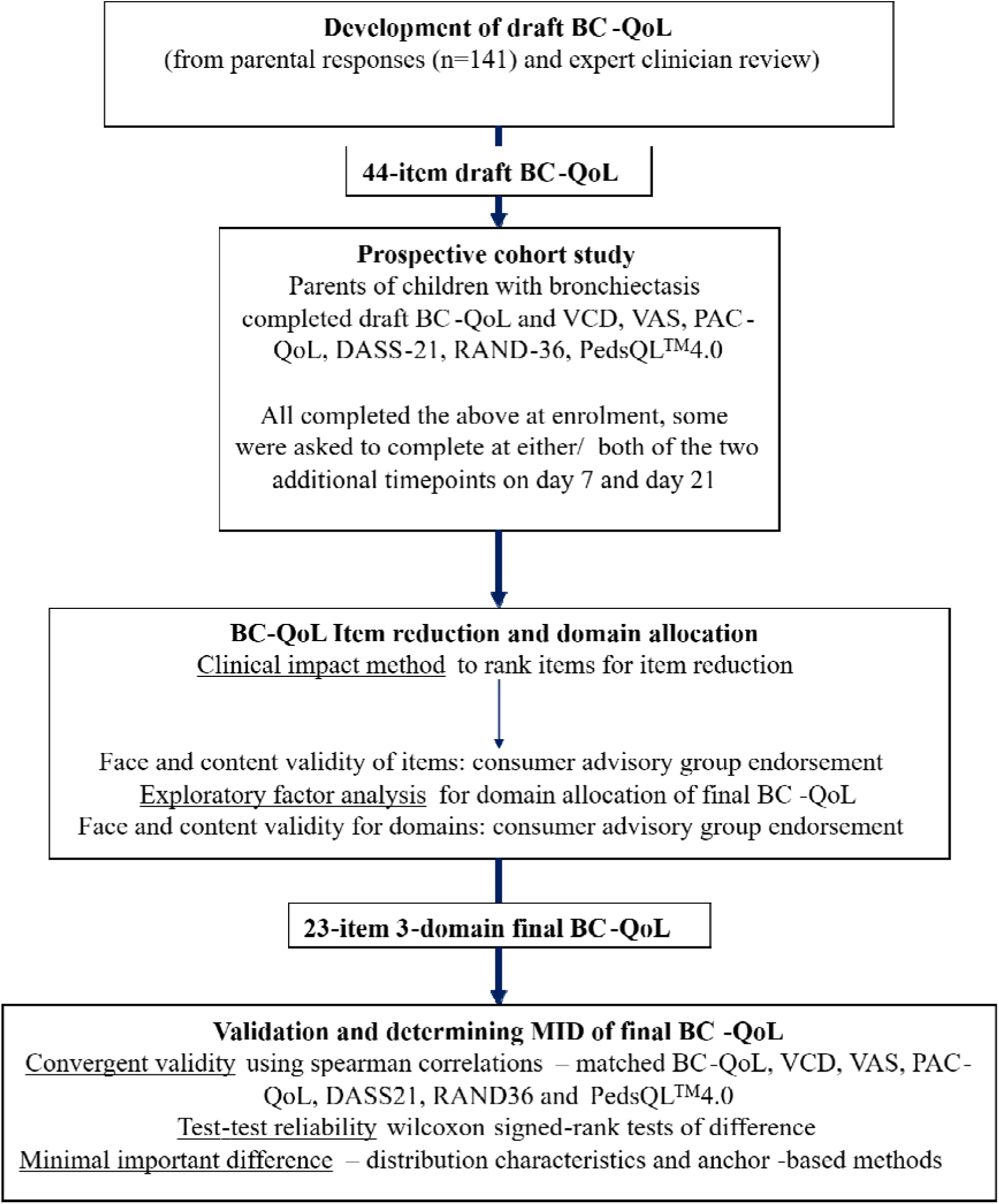
Study schematic. BC-QoL = bronchiectasis child-specific parent-proxy quality of life, MID = minimally important difference, PAC-QoL = parent acute cough quality of life questionnaire, VAS = visual analogue scale, VCD = verbal category descriptive, DASS-21 = Depression anxiety and stress scale, PedsQL^TM^4.0: paediatric quality of life index.

### Development of draft BC-QoL

We developed a 44-item draft BC-QoL with a 7-point Likert-type scale from: (a) responses from 142 parents to questions about the burden of bronchiectasis on their child, themselves, and their family;[4] (b) clinicians’ impression of parents’ burden; and (c) our previous experience developing child QoL instruments.[22,23] The next draft BC-QoL consisted of 16 items that reflected frequency of feelings and symptoms; “frequency items” and 28 reflecting parental worries and concerns; “worry items”.

We conducted a prospective cohort study at the Queensland Children’s Hospital, Brisbane, Australia. We enrolled parents of children with bronchiectasis who completed the draft BC-QoL and six other instruments at least once, at several timepoints during different bronchiectasis clinical states (stable, exacerbation, recovery) (Figure 2). Participants were required to complete these instruments at enrolment and were then asked to complete them at either or both of two additional timepoints, days 7 and day 21. The inclusion and exclusion criteria are described in eAppendix-1. We defined an *exacerbation* as a parent reported increase in frequency of wet cough for three or more days, with or without need for antibiotics. *Stable* state was defined as absence of an exacerbation in the preceding 4 weeks. We considered those who had recent (less than four weeks) resolution of an exacerbation to be in a *recovery* state.

**Figure 2:**
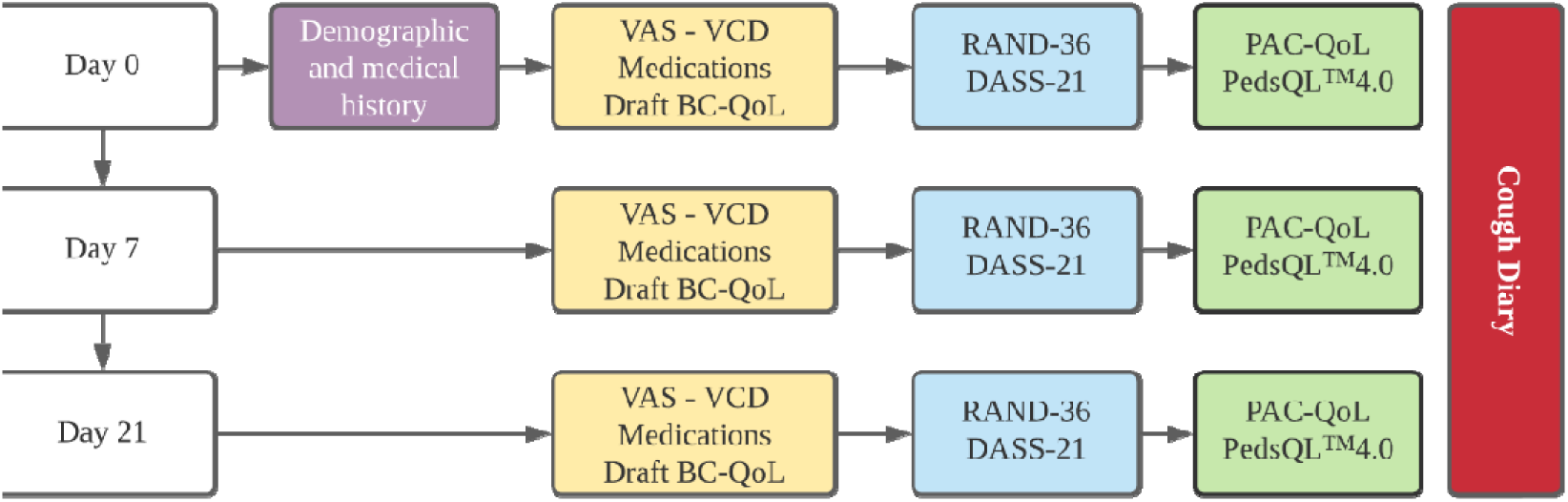
BC-QoL schematic of prospective cohort study procedures and materials. BC-QoL = bronchiectasis child-specific parent-proxy quality of life, VAS = visual analogue scale: cough severity, VCD = Verbal category descriptive cough score, DASS-21 = depression anxiety and stress scale 21 item, PAC-QoL = parent proxy acute cough quality of life, DASS-21 = Depression anxiety and stress scale, PedsQL^TM^4.0: paediatric quality of life index.

The six other instruments were used to assess convergent and discriminant validity of the final BC-QoL. All are validated instruments and consisted of: cough verbal category descriptive (VCD)[25] and cough visual analogue score (VAS),[26] parent proxy children’s acute cough specific QoL instrument (PAC-QoL),[22] depression, anxiety and stress 21-item scale (DASS-21),[27] RAND-36,[28] and paediatric quality-of-life inventory (PedsQL^TM^4.0).[29] VCD and VAS are validated measures of cough severity in children,[25,26] where higher scores indicate more severe cough. VCD is rated from 0 (no cough) to 5 (distressing cough) and the VAS ranged from 1 (no cough) to 10 (most severe cough). The PAC-QoL is a QoL measure specific for children with acute cough and is sensitive to changes in acute cough severity over short periods, between 3 and 14 days.[22] PedsQL^TM^4.0, a generic paediatric HR-QoL that measures overall health and well-being of children across four domains; physical, emotional, social and school functioning.[29] RAND-36 measures adult health across 8 domains: “physical functioning, role limitations caused by physical health problems, role limitations caused by emotional problems, social functioning, emotional well-being, energy/fatigue, pain, and general health perceptions”.[28] DASS-21 is a 21-item self-reporting scale for adults that measures depression, anxiety and stress. For both these measures, higher scores reflect greater severity and were used to measure parental health and mental status.[27] Justification of the use of these measures is further discussed in eAppendix-1.

### BC-QoL item reduction and domain allocation

We analysed responses to the draft BC-QoL using the clinical impact method of item reduction to rank the most important items,[15,23,30] and reduce the number of items, as described previously. [22,24] To assess the face and content validity of included items we presented the reduced item BC-QoL to our consumer advisory group, consisting of parents of children with bronchiectasis and/or young adults with bronchiectasis, who were able to endorse or remove items leading to a final BC-QoL. Next, we conducted exploratory factor analysis of the final BC-QoL to guide allocation of items to domains of physical, social and emotional based on the World Health Organization’s definition of health,[31] and as we have done previously.[22,24] We drafted item allocation to the 3 domains which was then presented to the consumer advisory group, who discussed the face and content validity of the domains, and suggested modifications.

### Validation and determining minimal important difference (MID) of final BC-QOL

We calculated the Cronbach’s alpha[32] and split half reliability adjusted with the Spearman-Brown prophecy formula[33] for the final BC-QoL. The medians for each domain and the total score for the BC-QoL were calculated and compared to each other and total scores of other instruments. To compare convergent validity, Spearman correlations were calculated between matched BC-QoL domain and total scores, VCD and VAS, and total and domain scores for PAC-QoL, DASS-21, RAND-36 and PedsQL^TM^4.0. We also explored the test-retest reliability of the final BC-QoL instrument using Wilcoxon signed-rank tests of difference for paired longitudinal responses within groups, and Wilcoxon rank-sum tests for unpaired responses between groups (see eAppendix-1). Finally, we calculated the minimal important difference for the final BC-QoL using two methods: distribution characteristics and anchor-based.[22,34] (see eAppendix-1).

## Results

### Demographics of prospective cohort study

We recruited 142 parents in our prospective cohort study. **Error! Reference source not found.** describes participant characteristics. The median (IQR) age of child participants was 5.9 years (IQR 3.5, 9.3), 85 (60%) were male, and 129 (91%) of parental respondents were female. All respondents completed the BC-QoL at least once, and most also completed the PedsQL^TM^4.0, DASS-21, RAND-36 and the PAC-QoL (**Error! Reference source not found.**). 101 participants completed at least one additional follow up time-point (Figure 3), and in total, we had 103 responses from parents of children in a stable state.

**Figure 3:**
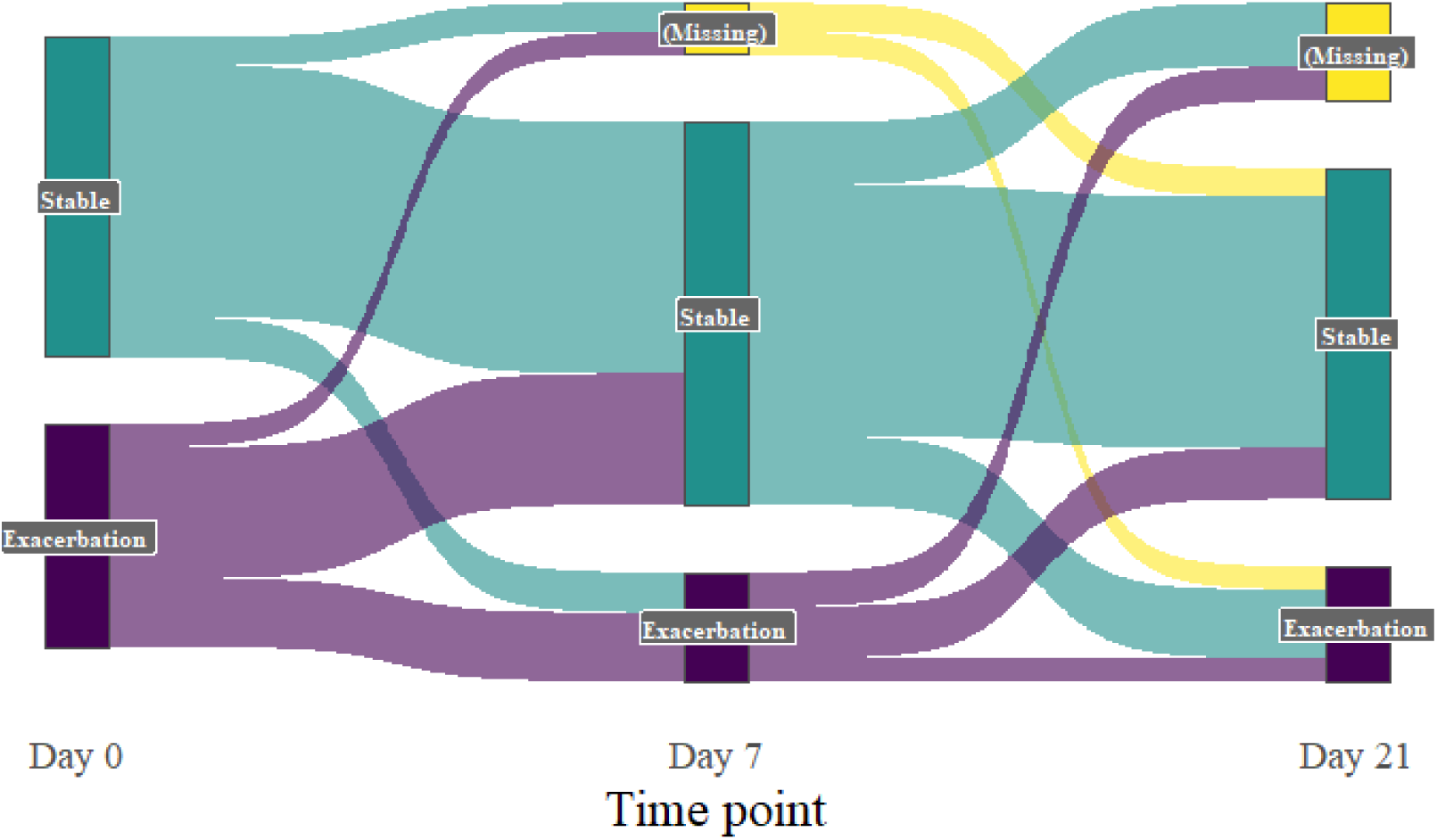
Sankey plot of participant exacerbation status over the longitudinal study (n= 101). Note, participants moving from exacerbated status to stable status are classified as recovered.

### Developing the final BC-QoL

The clinical impact analysis of stable state responses (n=103) indicated that the 21st item “How worried or concerned were you about your child’s bronchiectasis becoming worse/progressing?” which had an impact score of 1.40, was a suitable “natural break” in impact scores, and was our cut off for item selection. At this item, participant endorsement, i.e., a score ranked higher than, “Once in a while” for frequency items or “A little worried or concerned” for worry items was 50.5%. Endorsement for all items with higher impact scores was at least 48.5% (e Table 1). Several items had high item-item correlations, above 0.8 (e Table 2).

**Table 1:** Clinical and demographic characteristics.

| Characteristic<br>Median (IQR) or n (%) | Overall<br>N = 142 | Stable<br>N = 94 | Exacerbation<br>N = 48 |
| --- | --- | --- | --- |
| Age (years) | 5.9 (3.5, 9.3) | 7.2 (4.4, 10.6) | 4.1 (2.9, 7.3) |
| Child sex (%Male) | 85: (60%) | 55 (59%) | 30 (63%) |
| Respondent sex (%Female) | 129 (91%) | 84 (89%) | 45 (94%) |
| Exacerbations in the previous 12 months | 3 (2, 6) | 2.5 (2, 4) | 4.5 (2, 8) |
| Hospital admissions in prior 3 years | 1 (0, 2) | 0 (0, 1) | 1 (0, 2) |
| Aboriginal/ Torres Strait Islander Status | 19 (13%) | 9 (10%) | 10 (21%) |
| Cough at baseline |  |  |  |
| VCD daytime cough score | 1 (0, 3) | 1 (0, 2) | 3 (1, 4) |
| Missing | 7 | 3 | 4 |
| VAS cough score | 2 (1, 4) | 1 (1, 2) | 4 (2, 6) |
| Missing | 5 | 4 | 1 |
| Wet cough present | 76 (55%) | 34 (37%) | 42 (89%) |
| Missing | 4 | 3 | 1 |

IQR = interquartile range, VAS = Visual analogue scale, VCD = Verbal Category Descriptive

**Table 2:**
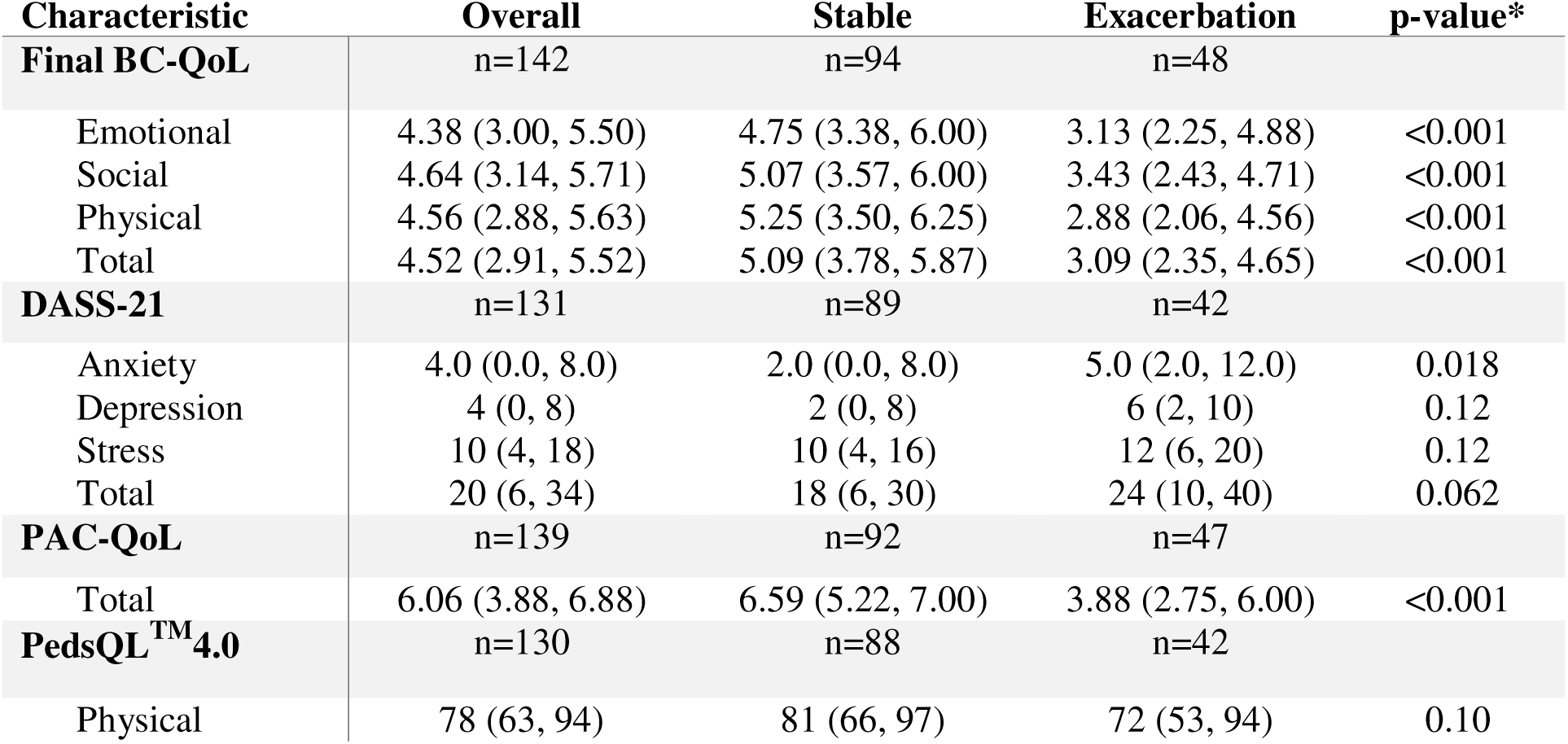
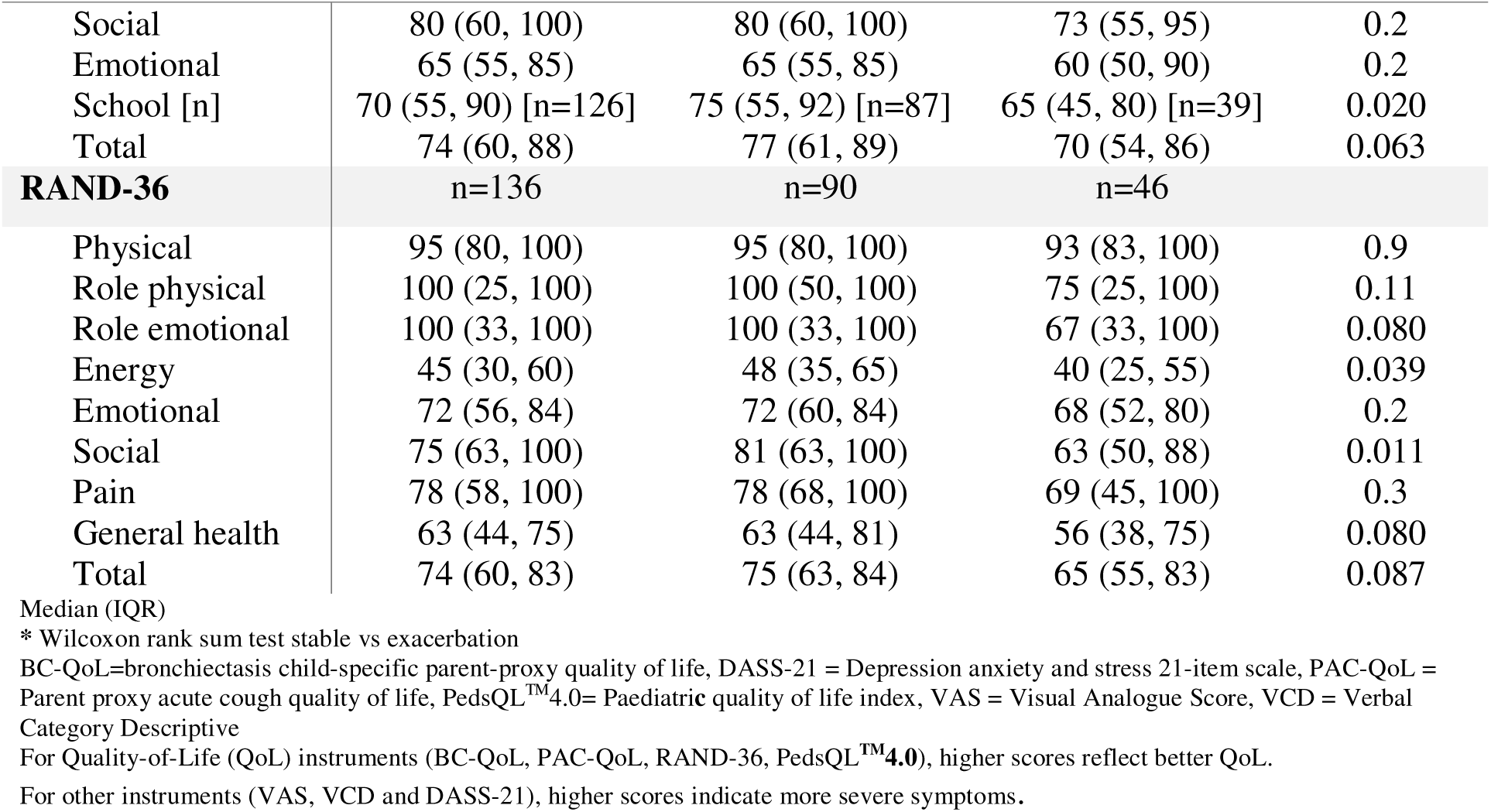
Responses to parent completed questionnaires and the final 23-item BC-QoL at enrolment.

Presentation of the clinical impact analysis (highest ranked 21 items for inclusion and remainder for removal) to the consumer advisory group saw three items recommended for re-inclusion to the instrument (items reflecting: parental concerns about doctors, feeling judged, and being awakened at night). One initially included item (energy) was recommended for removal by the group. The resulting final BC-QoL instrument after consumer advisory group endorsement consisted of 23 items.

The exploratory factor analysis of the final BC-QoL indicated a domain structure of 3 underlying factors (eTables 3 and 4). We presented this draft domain structure to the consumer advisory group, who discussed the face validity of this domain structure and applied their recommendations. The final modified domain structure included: eight items in the emotional and physical domains and seven in the social domain (eTable-1).

### Validity, reliability and responsiveness of the final BC-QoL

The median impact scores of items in the final BC-QoL in the stable enrolment group was 1.54 (IQR 1.42, 2.16) (eTable-1). No item had high rates of missing or declined responses. Inter-item correlations ranged from 0.28 to 0.84 and item-total correlations ranged from 0.53 to 0.89 (eTable-2). At both enrolment and day 21 follow-up, the final BC-QoL and each of its domains had high internal consistency (minimum values across all domains and timepoints: Cronbach’s α=0.90, split half reliability raw=0.85, adjusted=0.97) (Table 3). Additionally, each BC-QoL domain sub-score (emotional, social, physical) had high correlation with total and other domain scores (Table 3).

**Table 3:** Spearman correlations between final 23 item BC-QoL scores and other psychometric instruments between enrolment (T1) and final follow-up (T2), n=101.

|  | Total |  | Emotional |  | Social |  | Physical |  |
| --- | --- | --- | --- | --- | --- | --- | --- | --- |
|  | T1<sup>†</sup> | T2<sup>‡</sup> | T1<sup>†</sup> | T2<sup>‡</sup> | T1<sup>†</sup> | T2<sup>‡</sup> | T1<sup>†</sup> | T2<sup>‡</sup> |
| **BC-QoL** |  |  |  |  |  |  |  |  |
| Emotional | 0.94** | 0.97** | — | — | 0.84** | 0.92** | 0.84** | 0.93** |
| Social | 0.95** | 0.97** | 0.84** | 0.92** | — | — | 0.87** | 0.93** |
| Physical | 0.95** | 0.98** | 0.84** | 0.93** | 0.87** | 0.93** | — | — |
| Total | — | — | 0.94** | 0.97** | 0.95** | 0.97** | 0.95** | 0.98** |
| **Cough Instruments** |  |  |  |  |  |  |  |  |
| VCD | -0.44** | -0.60** | -0.40** | -0.62** | -0.38** | -0.56** | -0.49** | -0.61** |
| VAS | -0.51** | -0.68** | -0.45** | -0.69** | -0.45** | -0.61** | -0.57** | -0.69** |
| **DASS-21** |  |  |  |  |  |  |  |  |
| Anxiety | -0.46** | -0.39** | -0.43** | -0.37** | -0.43** | -0.36** | -0.45** | -0.35** |
| Depression | -0.34** | -0.43** | -0.37** | -0.42** | -0.31** | -0.44** | -0.31** | -0.39** |
| Stress | -0.39** | -0.37** | -0.39** | -0.36** | -0.35** | -0.35** | -0.37** | -0.34** |
| Total | -0.44** | -0.44** | -0.44** | -0.44** | -0.39** | -0.43** | -0.40** | -0.41** |
| **PAC-QoL** |  |  |  |  |  |  |  |  |
| Total | 0.77** | 0.83** | 0.72** | 0.81** | 0.71** | 0.79** | 0.78** | 0.83** |
| **PedsQL™4.0** |  |  |  |  |  |  |  |  |
| Physical | 0.39** | 0.49** | 0.38** | 0.44** | 0.43** | 0.49** | **0.33\*\*** | **0.47\*\*** |
| Social | 0.35** | 0.36** | 0.36** | 0.32** | **0.41\*\*** | **0.39\*\*** | 0.27** | 0.35** |
| Emotional | 0.43** | 0.52** | **0.43\*\*** | **0.46\*\*** | 0.43** | 0.52** | 0.39** | 0.50** |
| School function | 0.31** | 0.39** | 0.29** | 0.35** | 0.33** | 0.43** | 0.27** | 0.37** |
| Total | 0.44** | 0.52** | 0.43** | 0.46** | 0.48** | 0.55** | 0.37** | 0.51** |
| **RAND-36** |  |  |  |  |  |  |  |  |
| Physical | 0.10 | 0.07 | 0.05 | 0.04 | 0.13 | 0.10 | 0.11 | 0.06 |
| Role physical | 0.24** | 0.15 | 0.23** | 0.17 | 0.24** | 0.15 | 0.25** | 0.11 |
| Role emotional | 0.37** | 0.21 | 0.38** | 0.23* | 0.33** | 0.20 | 0.35** | 0.16 |
| Energy | 0.35** | 0.39** | 0.36** | 0.37** | 0.29** | 0.38** | 0.37** | 0.39** |
| Emotional | 0.41** | 0.37** | **0.43\*\*** | **0.40\*\*** | 0.35** | 0.36** | 0.38** | 0.33** |
| Social | 0.39** | 0.37** | 0.39** | 0.39** | **0.38\*\*** | **0.40\*\*** | 0.36** | 0.33** |
| Pain | 0.22* | 0.05 | 0.17* | 0.08 | 0.24** | 0.06 | 0.22* | 0.03 |
| General | 0.16 | 0.26* | 0.13 | 0.24* | 0.14 | 0.27* | 0.20* | 0.24* |
| Total | 0.33** | 0.32** | 0.31** | 0.33** | 0.31** | 0.33** | 0.33** | 0.26* |
| **Reliability indices** |  |  |  |  |  |  |  |  |
| Cronbach's $\alpha$ | 0.97 | 0.98 | 0.93 | 0.96 | 0.90 | 0.93 | 0.93 | 0.94 |
| SHR | 0.95 | 0.95 | 0.91 | 0.90 | 0.85 | 0.89 | 0.90 | 0.91 |
| SHR adjusted | 0.97 | 0.98 | 0.95 | 0.95 | 0.92 | 0.94 | 0.95 | 0.95 |

Correlations of related domains in **bold**. \* Significant at 5% level, \*\* Significant at the 1% level
<sup>†</sup>Time-point 1: Enrolment, <sup>‡</sup>Time-point 2: day 21 follow-up. SHR = Split-half reliability
BC-QoL = bronchiectasis child-specific parent-proxy quality of life, DASS-21 = Depression anxiety and stress 21-item scale, PAC-QoL = Parent proxy acute cough quality of life, PedsQL™4.0 = Paediatric quality of life index, VAS = Visual Analogue Score, VCD = Verbal Category Descriptive
For Quality-of-Life (QoL) instruments (BC-QoL, PAC-QoL, RAND-36, PedsQL™4.0), higher scores reflect better QoL.
For VAS, VCD and DASS-21, higher scores indicate more severe symptoms.

In evaluating how the BC-QoL correlates with other instruments to assess the convergent and discriminant validity of the final BC-QoL instrument (Table 3), we found that BC-QoL physical and emotional domain scores had high correlations with matching domains from both the RAND-36 and PedsQL^TM^4.0. BC-QoL domain and total scores had high correlations with PAC-QoL, VCD and VAS scores, particularly the physical domain, indicating sensitivity against changes in cough severity.

BC-QoL demonstrated evidence of good repeatability with high test-retest reliability between enrolment and follow-up in those whose exacerbation status was the same between both time-points (i.e. stable both at an enrolment and follow-up or exacerbation at both enrolment and follow-up) (n=62); the intraclass coefficient was 0.74 (95%CI 0.62-0.82). However, there were statistically significant but small magnitude changes in participants who were in a stable state at both enrolment and day 21.

The instrument demonstrated mixed performance when median changes over time were analysed between exacerbation status groups (Figure 4). BC-QoL scores in those who were stable at enrolment but had a new exacerbation at day 21 (n=14) decreased overall, however not all of these changes reached statistical significance. The instrument performed as expected in both groups who were in an exacerbation state at enrolment, with BC-QoL domain and total scores increasing in participants who had recovered from an exacerbation by day 21, and were essentially unchanged in those who reported remaining in an exacerbation state at day 21 (Figure 4).

**Figure 4:**
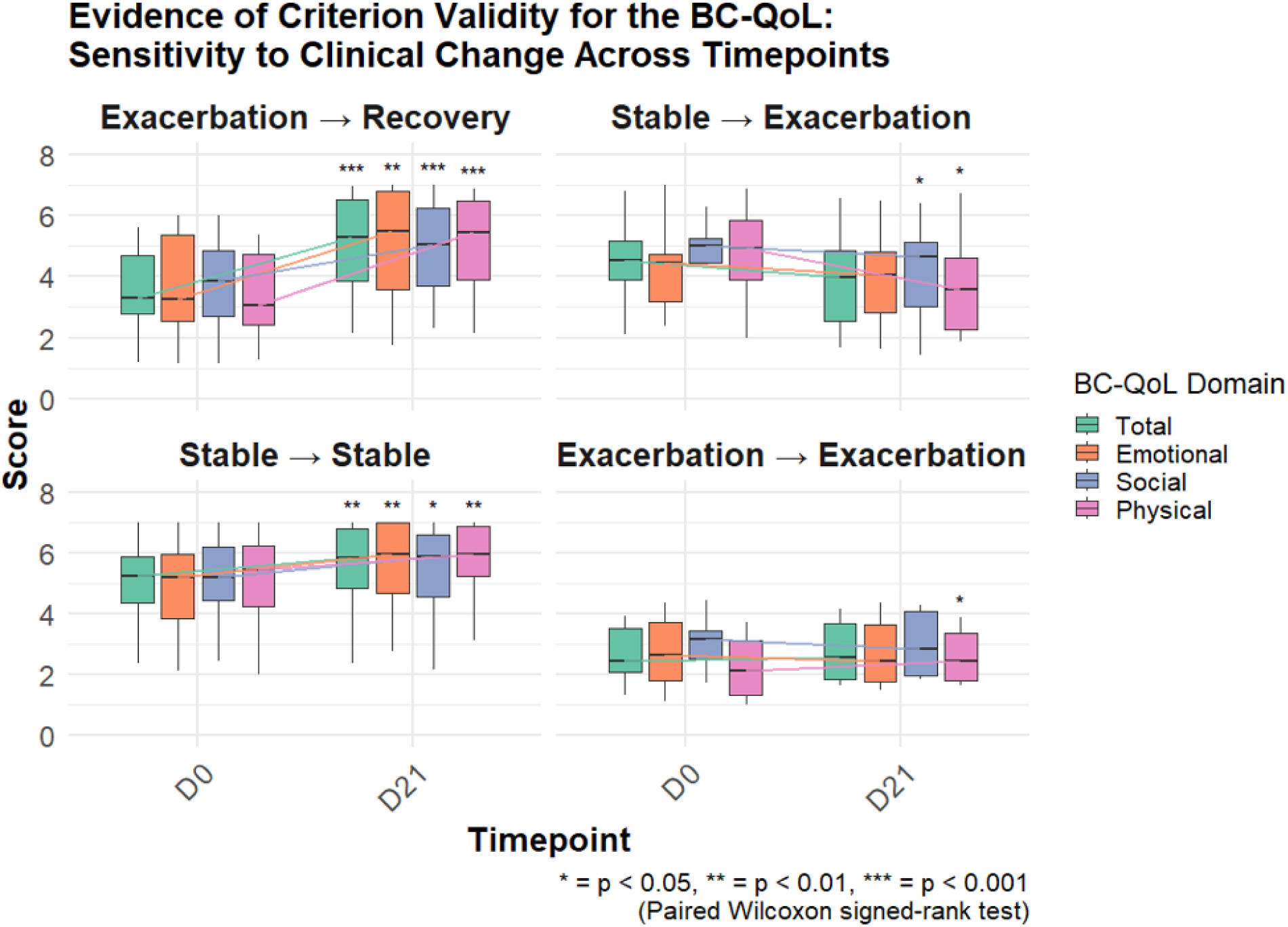
Paired responses between BC-QoL scores within groups between enrolment (D0) and follow-up (D21), grouped by exacerbation status.

Finally, when grouped by enrolment exacerbation status, no differences were detected in all total and domain scores at enrolment when their bronchiectasis clinical state was the same, but significant differences in follow-up scores were detected based on follow-up exacerbation status (Table 4).

**Table 4:**
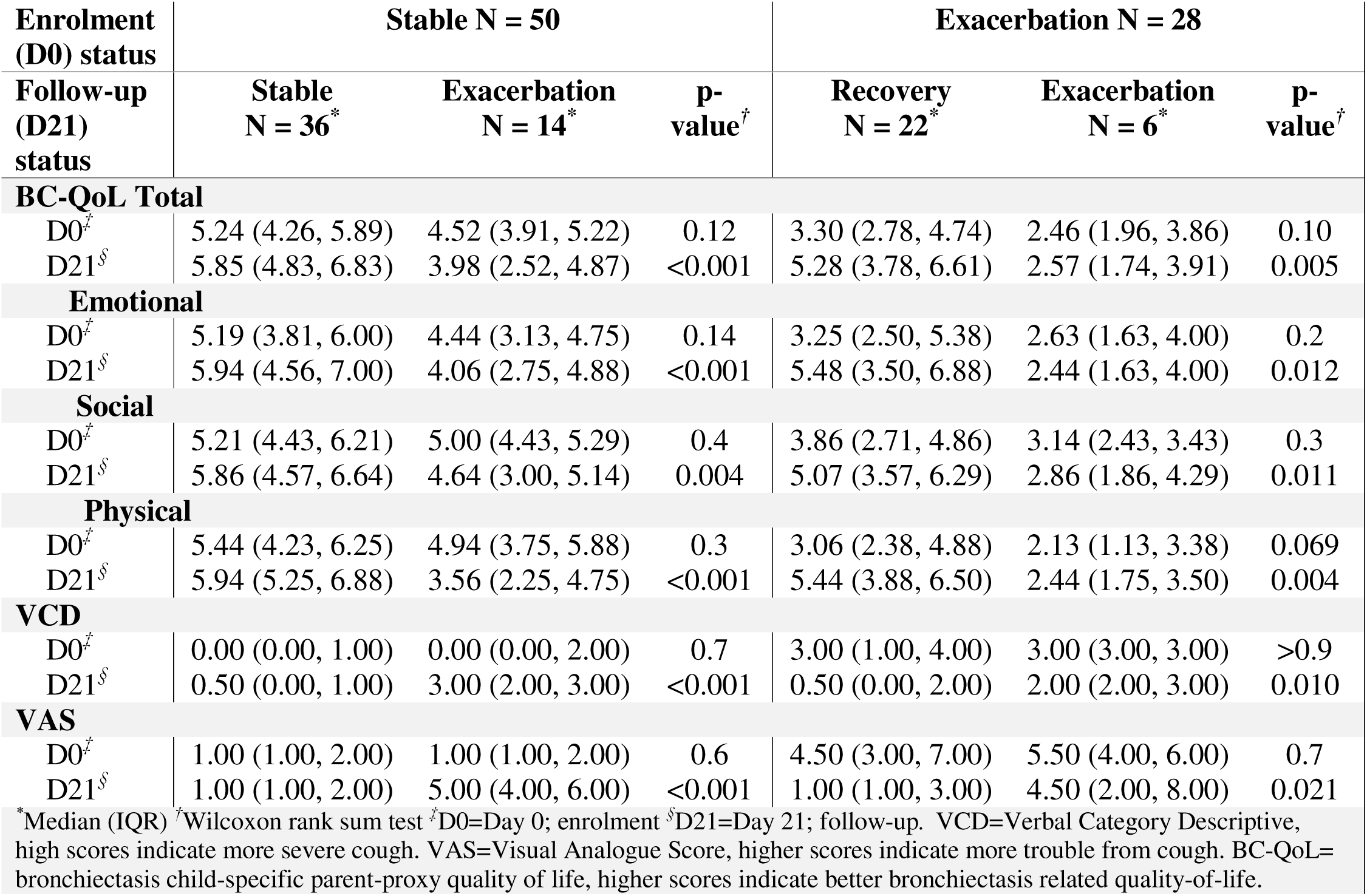
Unpaired responses in BC-QoL scores between enrolment (D0) and follow-up (D21) grouped by enrolment exacerbation status.

| Enrolment (D0) status | Stable N = 50 |  |  | Exacerbation N = 28 |  |  |
| --- | --- | --- | --- | --- | --- | --- |
| Follow-up (D21) status | Stable N = 36* | Exacerbation N = 14* | p-value<sup>‡</sup> | Recovery N = 22* | Exacerbation N = 6* | p-value<sup>‡</sup> |
| **BC-QoL Total** |  |  |  |  |  |  |
| D0<sup>‡</sup> | 5.24 (4.26, 5.89) | 4.52 (3.91, 5.22) | 0.12 | 3.30 (2.78, 4.74) | 2.46 (1.96, 3.86) | 0.10 |
| D21<sup>§</sup> | 5.85 (4.83, 6.83) | 3.98 (2.52, 4.87) | <0.001 | 5.28 (3.78, 6.61) | 2.57 (1.74, 3.91) | 0.005 |
| **Emotional** |  |  |  |  |  |  |
| D0<sup>‡</sup> | 5.19 (3.81, 6.00) | 4.44 (3.13, 4.75) | 0.14 | 3.25 (2.50, 5.38) | 2.63 (1.63, 4.00) | 0.2 |
| D21<sup>§</sup> | 5.94 (4.56, 7.00) | 4.06 (2.75, 4.88) | <0.001 | 5.48 (3.50, 6.88) | 2.44 (1.63, 4.00) | 0.012 |
| **Social** |  |  |  |  |  |  |
| D0<sup>‡</sup> | 5.21 (4.43, 6.21) | 5.00 (4.43, 5.29) | 0.4 | 3.86 (2.71, 4.86) | 3.14 (2.43, 3.43) | 0.3 |
| D21<sup>§</sup> | 5.86 (4.57, 6.64) | 4.64 (3.00, 5.14) | 0.004 | 5.07 (3.57, 6.29) | 2.86 (1.86, 4.29) | 0.011 |
| **Physical** |  |  |  |  |  |  |
| D0<sup>‡</sup> | 5.44 (4.23, 6.25) | 4.94 (3.75, 5.88) | 0.3 | 3.06 (2.38, 4.88) | 2.13 (1.13, 3.38) | 0.069 |
| D21<sup>§</sup> | 5.94 (5.25, 6.88) | 3.56 (2.25, 4.75) | <0.001 | 5.44 (3.88, 6.50) | 2.44 (1.75, 3.50) | 0.004 |
| **VCD** |  |  |  |  |  |  |
| D0<sup>‡</sup> | 0.00 (0.00, 1.00) | 0.00 (0.00, 2.00) | 0.7 | 3.00 (1.00, 4.00) | 3.00 (3.00, 3.00) | >0.9 |
| D21<sup>§</sup> | 0.50 (0.00, 1.00) | 3.00 (2.00, 3.00) | <0.001 | 0.50 (0.00, 2.00) | 2.00 (2.00, 3.00) | 0.010 |
| **VAS** |  |  |  |  |  |  |
| D0<sup>‡</sup> | 1.00 (1.00, 2.00) | 1.00 (1.00, 2.00) | 0.6 | 4.50 (3.00, 7.00) | 5.50 (4.00, 6.00) | 0.7 |
| D21<sup>§</sup> | 1.00 (1.00, 2.00) | 5.00 (4.00, 6.00) | <0.001 | 1.00 (1.00, 3.00) | 4.50 (2.00, 8.00) | 0.021 |
| *Median (IQR) <sup>‡</sup>Wilcoxon rank sum test <sup>‡</sup>D0=Day 0; enrolment <sup>§</sup>D21=Day 21; follow-up. VCD=Verbal Category Descriptive, high scores indicate more severe cough. VAS=Visual Analogue Score, higher scores indicate more trouble from cough. BC-QoL=bronchiectasis child-specific parent-proxy quality of life, higher scores indicate better bronchiectasis related quality-of-life. |  |  |  |  |  |  |

### Minimal important difference of the final BC-QoL

Using the distribution method, minimal important difference values ranged from 0.26 (based on the standard error of measurement) to 0.74 (based one half of the standard deviation) for the total scale and 0.42-0.85 across the domains (eTable-5). Using the anchor-based approach (see eAppendix-1), the minimal important difference for the total scores was 1.32 and domain scores ranged from 1.29 to 1.44. Overall, we therefore estimated that the minimal important difference for the total BC-QoL score lies between 0.74 and 1.32.

## Discussion

We have developed the first bronchiectasis child-specific parent-proxy quality-of-life (BC-QoL) instrument designed for use by parents of children with bronchiectasis and have provided evidence of its validity, reliability and responsiveness. The final 23 item three domain instrument, informed by exploratory factor analysis and the clinical impact method, was endorsed by a consumer advisory panel. We have also estimated the instrument’s MID to be between 0.74 and 1.32. The BC-QoL would be a valuable new patient-reported outcome measure to aid in understanding the burden of paediatric bronchiectasis on children and families and for use in intervention studies.

In adult bronchiectasis research and healthcare, condition specific measures of HR-QoL are available, namely the QoL-B[20] and the BHQ[21] These PROMs, shown to be useful outcome measures, are now widely used in clinical trials. However, these adult measures are not applicable to paediatric patients due to the important differences in child and adult based QoL instruments in general[3,23,29] in addition to important differences in the impact, treatment and prognosis of paediatric and adult bronchiectasis.[1] Our newly developed BC-QoL now provides a disease-specific patient-reported outcome measure in paediatric bronchiectasis clinical trials where generic measures or the parent-proxy child-cough quality of life (PC-QoL), which focusses only on the symptom of cough, have historically been utilised[3] because of the absence of a child specific bronchiectasis QoL.

Of the 23-items in the final BC-QoL instrument, eight each were allocated to the emotional and physical domains and seven to the social domain. These domains correspond to the World Health Organization definition of health as “A state of physical, mental and social wellbeing…”.[31] and the domains we have previously used in other instruments we developed.[22,24] In contrast to the PAC-QoL for acute cough, where most items lie in the physical domain,[22] items in the BC-QoL are spread evenly between domains. This reflects the effect of bronchiectasis on children beyond the physical effects to its social and psychological impact.[4] Notably, the final parent-informed domain structure was different to the one estimated in the factor analysis, particularly with regard to the social domain, which is indicative of a domain structure with stronger content validity than an empirically derived one.

Evidence of the convergent validity of the BC-QoL is reflected in the high Spearman correlations between BC-QoL domains and related domains of the PedsQL^TM^4.0 and the RAND-36, and discriminant validity is demonstrated by the low, non-significant correlations of BC-QoL scores and physical domains of the parent-completed RAND-36. Additionally, we demonstrated that the instrument has high split half reliability and high test-retest reliability. The final BC-QoL instrument is also sensitive to change over time to reflect the health and wellbeing of children with bronchiectasis, and the MID in total score is between 0.74 and 1.32. However, when using anchor-based methods in estimating the MID when changes in VCD were largest (>1), changes in BC-QoL scores were approximately the same as those with no change in VCD scores. It is likely because the sizes of these groups (moderate to extreme change) were too small (each <10 children) to obtain stable estimates of BC-QoL change with VCD change, or, perhaps given that bronchiectasis symptomology is known to extend beyond cough,[35] VCD alone may not be a good anchor. Future studies that include both the BC-QoL measure and other measures of bronchiectasis severity could explore the anchor-based MID of the BC-QoL.

The BC-QoL is a parent-proxy measure because it is standard practice for parents to act as proxies for their child’s health as young children cannot verbally describe their concerns. Whilst this is a parent-proxy measure of the child’s health, several items relate to the well-being and QoL of the parent, and thus, to some extent, this instrument may also measure parental spillover effects.[36] Notably, some items particularly concerned with the parent’s experience for example, (how often) “were you awakened…”, “did you feel upset…”, “did you feel anxious”, “did you worry…”, and “did you feel sorry…” were included in the final BC-QoL. This spillover is however relevant for our instrument as it is known parental QoL can be affected by their child’s chronic medical condition.[37,38]

In each stage of development of the BC-QoL, we have accounted for the lived experience of children with bronchiectasis and their caregivers by including consumer feedback. It is increasingly recognised that research with, rather than about or for is necessary to ensure outcomes and benefits relevant to the patient group.[39] The involvement of consumers is increasingly seen as mandatory in the planning and design of research studies.[40] This is an important strength of our new BC-QoL instrument and contributes to the high face validity of the instrument.

This study has several important limitations. Firstly, most data collection was during the COVID-19 pandemic. During this period, there was heightened concern about cough and other respiratory symptoms which could have influenced the response patterns to the BC-QoL and the instruments used to examine its convergent and discriminant validity. Secondly, as the median age of the children in our cohort was young, BC-QoL’s applicability to older children is potentially limited. Indeed, a child-answered bronchiectasis QoL instrument is required for older children,[17] particularly the adolescent age-group when the self-reported impact of their bronchiectasis may be very different that those identified by their parents. Respondents also largely self-selected into the follow-up part of the study which may limit the generalisability of follow-up results, and we did not account for variation based on aetiology and underlying disorders Lastly, our prospective cohort study was conducted in one specialist centre in Australia, which may limit the generalizability of the instrument to other populations. However, our consumer advisory panel is based throughout Australia, including rural and remote locations, which is an important feature of our study and arguably improves the generalizability of our BC-QoL instrument. This limitation could be further mitigated by multicentre or international validation studies with translation/ cultural adaptation.

## Conclusion

Our newly developed BC-QoL responds to the need identified by children with bronchiectasis and their parents as a core-outcome measure.[9] BC-QoL is a specific, sensitive and valid patient-reported outcome measure that can be used in evaluation of bronchiectasis interventions in children. Future research should explore the performance of the BC-QoL in different contexts including other centres, (especially in other countries), and in a post-pandemic society. In addition, the development of a child-answered bronchiectasis QoL instrument for older children is an important future research step. Item response theory techniques may be useful in exploring the relative “difficulty” of items and calibrating them to provide more precise and informative summary scores. Finally, to facilitate the implementation of the BC-QoL in economic analyses in clinical research trials, it could be converted to a preference-based measure for measuring health utility.

## Author contributions

The study was conceived and designed by ABC and JMM. Material preparation was performed by JMR, ABC & JMM. Data collection was performed by JMR, JMM and ABC. Data analyses were performed by JMR, PAN, JMM and ABC. The first draft of the manuscript was prepared by JMR and all authors commented on previous versions of the manuscript. All authors read and approved the final manuscript.

## Ethics approval

Ethical approval for this study was obtained from the Children’s Health Queensland Human Research Ethics (HREC/20/QCHQ/64160) and administrative approval from Queensland University of Technology 2000000433.

### Abbreviations

BC-QoL: bronchiectasis child-specific parent-proxy quality of life
HR-QoL: health related quality of life
MID: minimally important difference
QoL: quality of life
PACQLQ: paediatric asthma caregiver’s quality of life questionnaire
PC-QoL: parent-proxy child chronic cough quality of life questionnaire
PAC-QoL: parent acute cough quality of life questionnaire
VAS: visual analogue scale
VCD: verbal category descriptive
PROM: patient-reported outcome measure.

## Supporting information

eAppendix-1

eAppendix-2

## Data Availability

 All data produced in the present study are available upon reasonable request to the authors

## References

1. Chang AB, Bush A, Grimwood K. Bronchiectasis in children: diagnosis and treatment. Lancet. 2018;392(10150):866–79.

2. Goyal V, Grimwood K, Marchant J, Masters IB, Chang AB. Pediatric bronchiectasis: No longer an orphan disease. Pediatr Pulmonol. 2016;51(5):450–69.

3. Nathan AM, de Bruyne JA, Eg KP, Thavagnanam S. Review: Quality of Life in Children with Non-cystic Fibrosis Bronchiectasis. Front Pediatr. 2017;5:84.

4. Marchant JM, Cook AL, Roberts J, Yerkovich ST, Goyal V, Arnold D, et al. Burden of Care for Children with Bronchiectasis from Parents/Carers Perspective. J Clin Med. 2021;10(24):5856.

5. Roberts JM, Goyal V, Kularatna S, Chang AB, Kapur N, Chalmers JD, et al. The Economic Burden of Bronchiectasis: A Systematic Review. Chest. 2023;164(6):1396–421.

6. Goyal V, Grimwood K, Ware RS, Byrnes CA, Morris PS, Masters IB, et al. Efficacy of oral amoxicillin-clavulanate or azithromycin for non-severe respiratory exacerbations in children with bronchiectasis (BEST-1): a multicentre, three-arm, double-blind, randomised placebo-controlled trial. Lancet Respir Med. 2019;7(9):791–801.

7. Jones T, O’Grady KF, Goyal V, Masters IB, McCallum G, Drovandi C, et al. Bronchiectasis - Exercise as Therapy (BREATH): rationale and study protocol for a multi-center randomized controlled trial. Trials. 2022;23(1):292.

8. Marchant JM, Chang AB, Schutz KL, Versteegh L, Cook A, Roberts J, et al. Utility of a personalised Bronchiectasis Action Management Plan (BAMP) for children with bronchiectasis: protocol for a multicentre, double-blind parallel, superiority randomised controlled trial. BMJ Open. 2021;11(12):e049007.

9. Chang AB, Boyd J, Bush A, Hill AT, Powell Z, Zacharasiewicz A, et al. A core outcome set for bronchiectasis in children and adolescents for use in clinical research: an international consensus study. Lancet Respir Med. 2024;12(1):78–88.

10. Wiebe S, Guyatt G, Weaver B, Matijevic S, Sidwell C. Comparative responsiveness of generic and specific quality-of-life instruments. J Clin Epidemiol. 2003;56(1):52–60.

11. Peters M, Crocker H. Condition-specific measure. Encyclopedia of quality of life and well-being research: Springer; 2020. p. 1-3.

12. Hvidøre Study Group on Childhood Diabetes, Skinner T, Hoey H, McGee H, Skovlund S. A short form of the Diabetes Quality of Life for Youth questionnaire: exploratory and confirmatory analysis in a sample of 2,077 young people with type 1 diabetes mellitus. Diabetologia. 2006;49:621–8.

13. Waters E, Davis E, Mackinnon A, Boyd R, Graham HK, Kai Lo S, et al. Psychometric properties of the quality of life questionnaire for children with CP. Dev Med Child Neurol. 2007;49(1):49–55.

14. Marino BS, Tomlinson RS, Wernovsky G, Drotar D, Newburger JW, Mahony L, et al. Validation of the pediatric cardiac quality of life inventory. Pediatrics. 2010;126(3):498–508.

15. Juniper EF, Guyatt GH, Feeny DH, Ferrie PJ, Griffith LE, Townsend M. Measuring quality of life in the parents of children with asthma. Qual Life Res. 1996;5(1):27–34.

16. Newcombe PA, Sheffield JK, Chang AB. Minimally important change in a Parent-Proxy Quality-of-Life questionnaire for pediatric chronic cough. Chest. 2011;139(3):576–80.

17. Newcombe PA, Sheffield JK, Petsky HL, Marchant JM, Willis C, Chang AB. A child chronic cough-specific quality of life measure: development and validation. Thorax. 2016;71(8):695–700.

18. Roberts JM, Chang AB, Goyal V, Kapur N, Marchant JM, McPhail SM, et al. Rasch validation of the short form (8 item) PC-QoL questionnaire and applicability of use as a health state classification system for a new preference-based measure. Qual Life Res. 2024;33(7):1893–903.

19. Anderson-James S, Newcombe PA, Marchant JM, Turner CT, Chang AB. Children’s Acute Cough-Specific Quality of Life: Revalidation and Development of a Short Form. Lung. 2021;199(5):527–34.

20. Quittner AL, O’Donnell AE, Salathe MA, Lewis SA, Li X, Montgomery AB, et al. Quality of Life Questionnaire-Bronchiectasis: final psychometric analyses and determination of minimal important difference scores. Thorax. 2015;70(1):12–20.

21. Spinou A, Siegert RJ, Guan WJ, Patel AS, Gosker HR, Lee KK, et al. The development and validation of the Bronchiectasis Health Questionnaire. Eur Respir J. 2017;49(5).

22. Anderson-James S, Newcombe PA, Marchant JM, O’Grady KA, Acworth JP, Stone DG, et al. An acute cough-specific quality-of-life questionnaire for children: Development and validation. J Allergy Clin Immunol. 2015;135(5):1179–85 e1-4.

23. Newcombe PA, Sheffield JK, Juniper EF, Marchant JM, Halsted RA, Masters IB, et al. Development of a parent-proxy quality-of-life chronic cough-specific questionnaire: clinical impact vs psychometric evaluations. Chest. 2008;133(2):386–95.

24. Newcombe PA, Sheffield JK, Juniper EF, Petsky HL, Willis C, Chang AB. Validation of a parent-proxy quality of life questionnaire for paediatric chronic cough (PC-QOL). Thorax. 2010;65(9):819–23.

25. Chang AB, Newman RG, Carlin JB, Phelan PD, Robertson CF. Subjective scoring of cough in children: parent-completed vs child-completed diary cards vs an objective method. Eur Respir J. 1998;11(2):462–6.

26. Boulet LP, Coeytaux RR, McCrory DC, French CT, Chang AB, Birring SS, et al. Tools for assessing outcomes in studies of chronic cough: CHEST guideline and expert panel report. Chest. 2015;147(3):804–14.

27. Henry JD, Crawford JR. The short-form version of the Depression Anxiety Stress Scales (DASS-21): construct validity and normative data in a large non-clinical sample. Br J Clin Psychol. 2005;44(Pt 2):227–39.

28. Hays RD, Morales LS. The RAND-36 measure of health-related quality of life. Ann Med. 2001;33(5):350–7.

29. Varni JW, Burwinkle TM, Seid M, Skarr D. The PedsQL 4.0 as a pediatric population health measure: feasibility, reliability, and validity. Ambul Pediatr. 2003;3(6):329–41.

30. Juniper EF, Guyatt GH, Streiner DL, King DR. Clinical impact versus factor analysis for quality of life questionnaire construction. Journal of clinical epidemiology. 1997;50(3):233–8.

31. World Health Organization. Constitution of the World Health Organization. Geneva: WHO; 1947.

32. Cronbach LJ. Coefficient alpha and the internal structure of tests. Psychometrika. 1951;16(3):297–334.

33. Spearman C. Correlation Calculated from Faulty Data. British Journal of Psychology, 1904-1920. 2011;3(3):271–95.

34. Yost KJ, Eton DT. Combining distribution- and anchor-based approaches to determine minimally important differences: the FACIT experience. Eval Health Prof. 2005;28(2):172–91.

35. Blamires J, Dickinson A, Byrnes CA, Tautolo ES. Sore and tired. A qualitative study exploring the symptom experience of youth with bronchiectasis. J Child Health Care. 2023;27(4):587–98.

36. Leech AA, Lin PJ, D’Cruz B, Parsons SK, Lavelle TA. Family Spillover Effects: Are Economic Evaluations Misrepresenting the Value of Healthcare Interventions to Society? Appl Health Econ Health Policy. 2023;21(1):5–10.

37. Smith S, Tallon M, Clark C, Jones L, Mörelius E. “You Never Exhale Fully Because You’re Not Sure What’s NEXT”: Parents’ experiences of stress caring for children with chronic conditions. Frontiers in pediatrics. 2022;10:902655.

38. Klassen AF, Klaassen R, Dix D, Pritchard S, Yanofsky R, O’Donnell M, et al. Impact of caring for a child with cancer on parents’ health-related quality of life. J Clin Oncol. 2008;26(36):5884–9.

39. Brett J, Staniszewska S, Mockford C, Herron Marx S, Hughes J, Tysall C, et al. Mapping the impact of patient and public involvement on health and social care research: a systematic review. Health expectations. 2014;17(5):637–50.

40. Australian Commission on Safety and Quality in Health Care. The National Clinical Trials Governance Framework and user guide for health service organisations conducting clinical trials. Sydney: ACSQHC; 2022.

