## Supplementary material for "Development and validation of a parent proxy bronchiectasis child quality of life instrument: The BC-QoL": eAppendix-1

### E-Appendix 1:

### Methods

#### Development of a 44-item draft BC-QoL

Items for the draft BC-QoL questionnaire were generated from 3 sources: (a) parental responses to questions about the burden of bronchiectasis on their child, themselves, and their family; (b) clinicians’ impression of parents’ burden; and (c) a previously developed child HR-QoL instrument.^1,2^ Data for part (a) were obtained from 141 parental free form responses to the question “what are 5 things that worry or concern you about your child’s bronchiectasis” collected in a previously reported study.^3^ Based on the development of previous HR-QoL instruments, we created items to match stems based on frequency (During the past week how often…) and worry/concern (During the past week how worried or concerned were you …). All items were rated on a 7 level Likert scale, ranging from 1 (All the time) to 7 (None of the time) for the frequency items and 1 (Very, very worried or concerned) to 7 (Not worried or concerned) for worry items. Higher scores therefore reflect better quality of life.

Before data collection began for the next phase, we presented the draft 44-item questionnaire to our advisory group of parents and children with bronchiectasis who had the opportunity to either add new items that they felt covered important QoL concerns, and/ or to remove irrelevant items.

#### Prospective cohort study

In the next phase, we conducted a prospective cohort study of the parents of children attending the respiratory outpatient department at the Queensland Children’s Hospital in Brisbane, Australia. Our study procedures were informed by previous studies conducted by ourselves and others in the development of other respiratory-specific QoL instruments.^1,2,4-8^ Participants and their children were approached either in person or by telephone and invited to participate in the study over one to three time points; Day 0, 7, and 21. They were not required to complete all three time-points.

Inclusion criteria were: (a) parents of children with respiratory physician diagnosed bronchiectasis (which was defined as mkn the presence of a clinical syndrome with abnormally dilated airways (bronchoarterial ratio [BAR] >0.8) detected on a multidetector computed tomography (MDCT) scan with cHRCT reconstruction),^9^ (b) child had at least two acute respiratory exacerbations in the twelve months before enrolment, and (c) parents were able to complete study procedures. Participants were excluded if they had a diagnosis of cystic fibrosis, severe cerebral palsy or any other severe neurodevelopmental condition.

##### Study materials and methods

After obtaining informed consent, demographic information about the parent and the child, and the child’s bronchiectasis and medical history were collected using a standardised case report form. The child’s current exacerbation status was elucidated on discussion with the parent and where required, with consultation with the child’s physician. At enrolment, parents could complete either a paper copy of the study instruments, including the draft 44 item BC-QoL, or on a tablet using the redcap mobile app, or at home online through redcap surveys.^10^ Parents also completed six other measures in the following order after completing the draft BC-QoL

##### Measures of cough severity

In addition to the draft BC-QoL, parents completed the cough verbal category descriptive (VCD) and cough visual analogue scores (VAS) which have been shown to be reflective of objective measures of cough frequency.^11^ In both measures, higher scores are indicative or more severe cough, where the VCD is rated from 0 (no cough) to 5 (distressing cough) and the VAS is rated from 1 – 10, where 1 is no trouble and 10 is very troublesome.

##### PAC-QoL

The parent proxy children’s acute cough specific QoL instrument (PAC-QoL) is a QoL measure specific for children experiencing acute cough with evidence supporting its validity and reliability.^1^ It consists of 16 items across three domains of physical, social and psychological wellbeing, and is sensitive to changes in acute cough severity over short periods, between 3 and 14 days.^1^

##### RAND-36

Parents also reported on their own health status by completing the RAND-36 which measures their own health across 8 domains: “physical functioning, role limitations caused by physical health problems, role limitations caused by emotional problems, social functioning, emotional well-being, energy/fatigue, pain, and general health perceptions”.^12^ Higher scores are reflective of worse health states.

##### DASS-21

Finally, because it is known that childhood bronchiectasis has a high psychological burden on parents,^13^ they completed the depression, anxiety and stress scale 21 item version (DASS-21).^14^

##### PedsQL^TM^4.0

The paediatric quality of life inventory (PedsQL^TM^4.0) is an instrument used for measuring general health and well-being of children across four domains, physical, emotional, social and school functioning.^15^ There are four versions specific for use in 2-4 year olds, 5-7 year olds, 8-12 year olds and 12 years and older. There is evidence supporting its validity and reliability and it can distinguish between children with and without chronic health conditions.^15^

#### Item reduction

Screening items for inclusion in the final questionnaire was done in stages. Responses to the 44-item draft questionnaire from participants in a stable state at enrolment were included for this analysis. First, for the clinical impact method only,^16^ each item was reverse coded so higher scores indicated worse quality of life, and descriptive statistics for each were calculated, including mean, median, missingness and ceiling/ floor effects of greater than 30% of responses. Secondly, clinical impact scores were calculated by multiplying the mean score of the reversed item by the proportion of respondents scoring the item higher than reversed level 3, “Once in a while” for frequency, “A little worried/ concerned” for worry items, as described elsewhere.^1,2,16^ We then ranked items based on clinical impact scores and searched for a natural break in scores, which we defined as a large difference in impact score between adjacent items, and used this break to define a cut-off for items we planned to retain. Frequency of endorsement and impact scores rankings for a group of children in an exacerbation state were also considered when determining the clinical impact cut off. Item-item and item-total Spearman’s rho correlations were calculated and items with high item-item correlation greater but the lower item-total correlation in the pair were recommended for removal.

#### Assessing content validity with the parent advisory group

The draft reduced-item BC-QoL was presented to an advisory group of 10 parents of children with bronchiectasis over two sessions, who discussed the face and contented validity of items, and who suggested the re-addition and removal of items from the scale based on this. Further details about the **Aus**tralian **Br**onchiectasis Centre of **R**esearch **E**xcellence especially for **A**boriginal and **T**orres Strait Islander C**h**ildr**e**n (AusBREATHE) Parent and Community Advisory Group (PAG) are available here: <https://www.crelungs.org.au/cre-parent-and-community-advisory-group>.

#### Domain allocation

To explore factorability, we calculated Kaiser-Meyer-Olkin measures of sampling adequacy, with a threshold of 0.8,^17^ and Bartlett’s test of sphericity.^18,19^ To guide the number of factors in exploratory factor analysis (EFA), we conducted Catell’s scree test including calculation of both eigenvalues and parallel analysis.^20^ EFA was conducted based on the polychoric correlation matrix of stable state responses. We estimated the model using principal axis method and oblimin rotation was applied to the factors due to correlations between them. This provisional empirically derived factor structure was presented again to the parent advisory group, who discussed the face and content validity of the domains, and suggested modifications.

#### Assessing final BC-QoL validity, reliability and responsiveness

We calculated the Cronbach’s alpha^21^ and split half reliability adjusted with the Spearman-Brown prophecy formula^22^ for the final BC-QoL. The medians for each domain and the total score for the reduced BC-QoL were calculated and compared to each other domain and total scores of other measures. To compare convergent validity, spearman correlations were calculated between matched BC-QoL domain and total scores, cough scores, and total and domain scores for PAC-QoL, DASS-21, RAND-36 and PedsQL^TM^4.0.

When considering test-retest reliability, it was important to first classify participants according to their exacerbation status at each assessment time-point to ensure only participants in comparable states of exacerbation. We divided respondents into four groups based on their exacerbation states at both enrolment and day 21 follow up, specifically: i) stable and stable, ii) stable and exacerbated, iii) exacerbated and recovery and iv) exacerbated and exacerbated, at enrolment and follow-up respectively. Wilcoxon signed-rank tests of difference were calculated for paired longitudinal responses within groups, and Wilcoxon rank-sum tests were calculated for unpaired responses between groups.

#### Calculation of minimal important difference of final BC-QoL

We used two approaches to calculate the minimal important differences in the final 23-item BC-QoL. Both methods necessitate the use of parametric summary statistics (means and standard deviations, SD).^1,23^ First, distribution characteristics were calculated by estimating the SD and transforming it to either standard error by multiplying it by the square root of 1 minus Cronbach’s alpha of the scale or dividing by one third and one half of the SD.^1^ For this study we calculated each but proceeded to use one half of the standard deviation method. Finally, the effect was calculated by taking the absolute difference of mean scores between the first and final time points and dividing them by the standard deviation of the scale at the first time-point. Anchor-based differences were estimated by calculating the absolute difference between enrolment and follow-up between BC-QoL and cough scores. Differences were grouped by changes in cough score and mean and standard deviations were estimated for each scale in that group.

#### Statistical analysis and sample size calculation

Based on the item reduction and validation of our chronic cough PC-QoL, a minimum of 60 children with complete data at Days 7 and 21 are required for analysis using the clinical impact method. For factor analysis, and given our experience performing this analysis on similar instruments, we expected to find approximately 3 factors, with between 15 and 30 items, which according to Mundform et al,^24^ could be adequately served by a sample of 85 participants, from which we conservatively aimed recruit at least 100 participants with stable enrolment responses.^24^

All analyses were performed in R.^25^ Descriptive statistics of participant characteristics were calculated using the gtsummary package,^26^ using non-parametric statistics because of non-normality in the data. We primarily estimated medians and interquartile ranges (IQR), and comparisons between groups were estimated using Pearson’s Chi-squared test for dichotomous variables, Fisher’s exact test for categorical variables, Wilcoxon’s rank sum test for unpaired continuous variables, and Wilcoxon’s signed rank test for paired continuous variables. The psych package was used for estimation of reliability statistics and exploratory factor analysis.^27^ The SimplyAgree package was used to explore test-retest validity.^28^

### Results

#### Calculation of minimally important difference

Using the anchor method demonstrated substantial absolute differences in BC-QoL scores between enrolment and follow-up for small changes in VCD compared to no change (0.82-1.32) (eTable 6). These differences in BC-QoL scores were not seen across larger changes in VCD scores, however, the number of respondents in these categories were notably small.

### Tables

eTable 1: Draft BC-QoL Item characteristics. Descriptive statistics, Impact score, and impact score ranks for items in cohort in stable state and exacerbated state.

| Item | **Code** | **Mean (SD)** *^‡^* | **Median (IQR)** *^‡^* | **Endorse** | **Impact score** | **Stable** **ranking** | **Exacerbated ranking** | **Domain** | **Density plot***^‡^* |
| --- | --- | --- | --- | --- | --- | --- | --- | --- | --- |
| *^*^* About your child being exposed to others who may be ill? | exposed | 4.18 (2.13) | 4 (3-6) | 74.8% | 3.12 | 1.00 | 7.00 | Social | 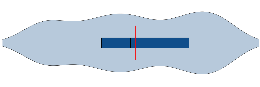 |
| *^†^* Did you feel overly protective toward your child because of his/her bronchiectasis? | overprotective | 3.99 (2.07) | 4 (2-6) | 70.9% | 2.83 | 2.00 | 8.00 | Emotional | 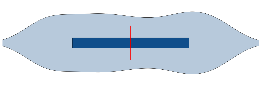 |
| *^*^* About long term damage to your child's lungs because of the bronchiectasis? | lungdamage | 3.81 (2.01) | 4 (2-5.5) | 72.8% | 2.77 | 3.00 | 4.00 | Physical | 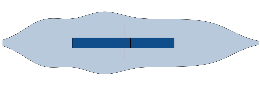 |
| *^†^* Did you feel sorry for your child because of his/her bronchiectasis? | sorry | 3.75 (2.17) | 4 (2-6) | 65% | 2.44 | 4.00 | 2.00 | Emotional | 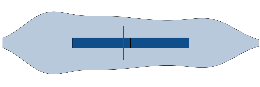 |
| *^*^* About the effects of your child's bronchiectasis on him/her? | effects | 3.45 (1.97) | 3 (2-5) | 63.1% | 2.17 | 5.00 | 9.00 | Social | 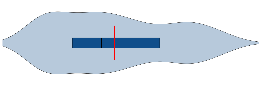 |
| *^†^* Did you worry about your child's bronchiectasis? | worry | 3.45 (1.94) | 3 (2-5) | 63.1% | 2.17 | 6.00 | 11.00 | Emotional | 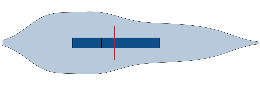 |
| *^†^* Did you feel anxious because of your child's bronchiectasis? | anxious | 3.40 (1.88) | 3 (2-5) | 63.1% | 2.14 | 7.00 | 10.00 | Emotional | 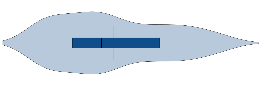 |
| *^*^* About your child feeling tired because of their bronchiectasis? | childtired | 3.32 (2.07) | 3 (1-5) | 58.3% | 1.94 | 8.00 | 14.00 | Emotional | 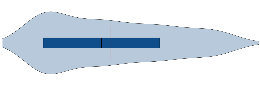 |
| *^*^* About your child feeling healthy? | healthy | 3.28 (1.97) | 3 (1-5) | 56.3% | 1.85 | 9.00 | 12.00 | Physical | 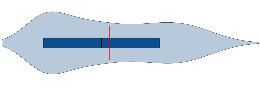 |
| *^*^* About your child taking too many antibiotics? | antibiotics | 3.29 (2.27) | 3 (1-5) | 52.4% | 1.73 | 10.00 | 6.00 | Physical | 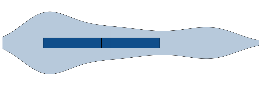 |
| *^*^* About your child's cough? | childscough | 2.97 (1.89) | 3 (1-4) | 52.4% | 1.56 | 11.00 | 1.00 | Physical | 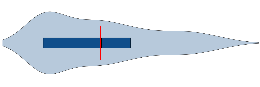 |
| *^*^* About your child not being able to play sport and exercise like other children? | sport | 2.99 (1.88) | 3 (1-4) | 51.5% | 1.54 | 12.00 | 31.00 | Social | 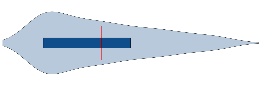 |
| *^*^* About your child's ability to lead a normal life as an adult? | normallife | 3.10 (2.02) | 2 (1-4) | 49.5% | 1.53 | 13.00 | 21.00 | Social | 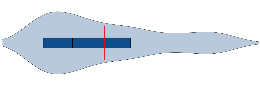 |
| *^*^* About your child's growth and weight? | growth | 3.08 (2.16) | 3 (1-5) | 49.5% | 1.52 | 14.00 | 29.00 | Physical | 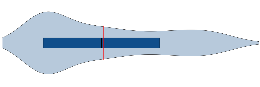 |
| *^*^* About your child feeling unwell because of his/her bronchiectasis? | unwell | 2.85 (1.75) | 3 (1-4) | 52.4% | 1.50 | 15.00 | 3.00 | Emotional | 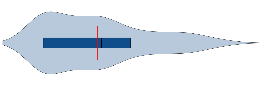 |
| *^†^* Did you feel upset because of your child's bronchiectasis? | upset | 2.99 (1.86) | 3 (1-5) | 49.5% | 1.48 | 16.00 | 17.00 | Emotional | 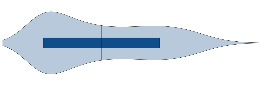 |
| *^*^* About your child having a lack of energy? | energy | 2.89 (1.91) | 3 (1-4) | 50.5% | 1.46 | 17.00 | 37.00 | ❌ | 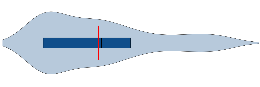 |
| *^*^* About your child being able to do normal activities such as playing and schoolwork because of his/her bronchiectasis? | activities | 2.72 (1.54) | 3 (1-4) | 52.4% | 1.42 | 18.00 | 18.00 | Social | 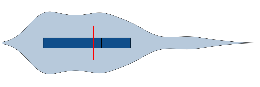 |
| *^*^* About your child not sleeping well because of the bronchiectasis? | sleep | 2.93 (1.96) | 2 (1-4) | 48.5% | 1.42 | 19.00 | 13.00 | Physical | 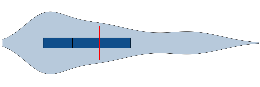 |
| *^*^* About your child's appetite? | appetite | 2.79 (1.86) | 3 (1-4) | 50.5% | 1.41 | 20.00 | 33.00 | Social | 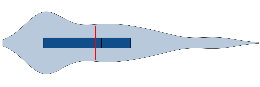 |
| *^*^* About your child's bronchiectasis becoming worse/progressing? | worse | 2.78 (1.69) | 3 (1-4) | 50.5% | 1.40 | 21.00 | 5.00 | Physical | 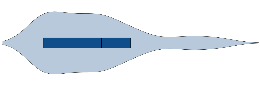 |
| *^*^* About your child having a shortened life span due to his/her bronchiectasis? | bronch | 2.99 (2.07) | 2 (1-4) | 45.6% | 1.36 | 22.00 | 27.00 | ❌ | 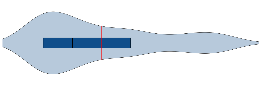 |
| *^†^* Did you feel down because of your child's bronchiectasis? | down | 2.70 (1.66) | 2 (1-4) | 49.5% | 1.34 | 23.00 | 25.00 | ❌ | 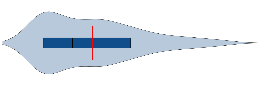 |
| *^*^* About your child being short of breath? | sob | 2.79 (1.79) | 2 (1-4) | 46.6% | 1.30 | 24.00 | 30.00 | ❌ | 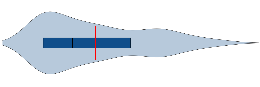 |
| *^*^* About the medication/s your child is taking for his/her bronchiectasis? | meds | 2.78 (1.90) | 2 (1-4) | 46.6% | 1.29 | 25.00 | 22.00 | ❌ | 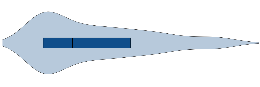 |
| *^†^* Were you awakened during the night because of your child's cough from his/her bronchiectasis? | awakened | 2.64 (1.76) | 2 (1-4) | 48.5% | 1.28 | 26.00 | 19.00 | Physical | 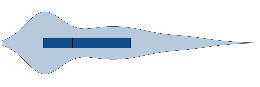 |
| *^*^* About not being able to do your child's treatments for bronchiectasis each day? | treatment | 2.61 (1.84) | 2 (1-4) | 43.7% | 1.14 | 27.00 | 36.00 | ❌ | 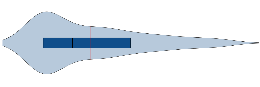 |
| *^*^* About your child's sputum or congestion? | sputum | 2.74 (1.98) | 2 (1-4) | 40.8% | 1.11 | 28.00 | 15.00 | ❌ | 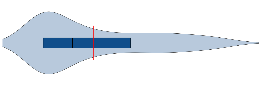 |
| *^†^* Did you feel tired or exhausted because of your child's bronchiectasis? | tired | 2.47 (1.66) | 2 (1-4) | 43.7% | 1.08 | 29.00 | 20.00 | ❌ | 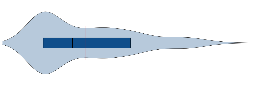 |
| *^*^* About your child's bronchiectasis symptoms bothering or being judged by other people? | judged | 2.72 (2.00) | 2 (1-4) | 38.8% | 1.06 | 30.00 | 16.00 | Social | 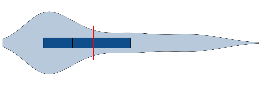 |
| *^†^* Did your child's bronchiectasis affect planning for your holidays? | holiday | 2.58 (1.94) | 2 (1-4) | 40.8% | 1.05 | 31.00 | 42.00 | ❌ | 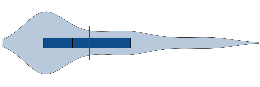 |
| *^†^* Did your child's bronchiectasis interfere with your job or work around the house? | work | 2.41 (1.68) | 2 (1-3) | 40.8% | 0.98 | 32.00 | 32.00 | ❌ | 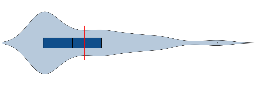 |
| *^†^* Were you concerned about not knowing enough about bronchiectasis to manage your child's condition? | notknowning | 2.50 (1.73) | 2 (1-4) | 38.8% | 0.97 | 33.00 | 38.00 | ❌ | 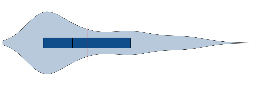 |
| *^†^* Were you or your family disturbed by your child's bronchiectasis? | disturbed | 2.40 (1.68) | 2 (1-3) | 39.8% | 0.95 | 34.00 | 24.00 | ❌ | 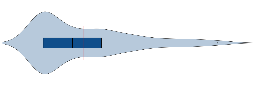 |
| *^*^* About your child missing school? | school | 2.62 (1.92) | 2 (1-4) | 35% | 0.92 | 35.00 | 26.00 | ❌ | 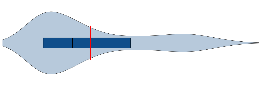 |
| *^*^* About your child getting enough physiotherapy for his/her bronchiectasis? | physio | 2.53 (1.88) | 2 (1-4) | 35.9% | 0.91 | 36.00 | 34.00 | ❌ | 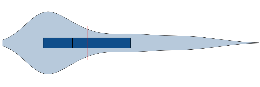 |
| *^†^* Were you financially burdened because of your child's bronchiectasis? | finance | 2.42 (1.86) | 1 (1-3) | 35.9% | 0.87 | 37.00 | 41.00 | ❌ | 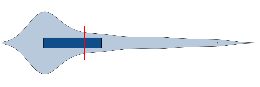 |
| *^*^* About your child feeling anxious because of his/her bronchiectasis? | childanxious | 2.53 (1.88) | 2 (1-4) | 34% | 0.86 | 38.00 | 39.00 | ❌ | 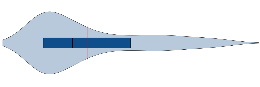 |
| *^†^* Did you worry about your child having to be admitted to hospital because of their bronchiectasis? | hospital | 2.45 (1.88) | 2 (1-3.5) | 35% | 0.85 | 39.00 | 28.00 | ❌ | 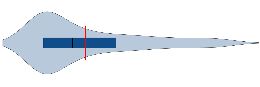 |
| *^*^* About your child having chest pain? | chest | 2.45 (1.89) | 2 (1-3) | 34% | 0.83 | 40.00 | 40.00 | ❌ | 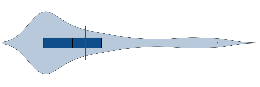 |
| *^*^* About when you should take your child to a doctor or emergency ward because of his/her bronchiectasis? | emergency | 2.39 (1.82) | 2 (1-4) | 31.1% | 0.74 | 41.00 | 23.00 | ❌ | 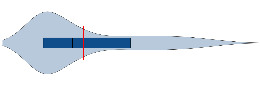 |
| *^†^* Did your family need to change plans because of your child's bronchiectasis? | plans | 2.25 (1.59) | 2 (1-4) | 31.1% | 0.70 | 42.00 | 35.00 | ❌ | 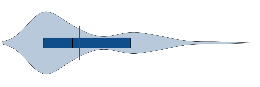 |
| *^†^* Were you concerned that doctors did not know how to treat your child's bronchiectasis? | doctors | 1.99 (1.62) | 1 (1-3) | 26.2% | 0.52 | 43.00 | 44.00 | Emotional | 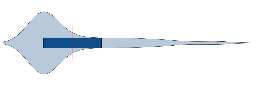 |
| *^*^* About your child choking? | choking | 2.10 (1.83) | 1 (1-2) | 23.3% | 0.49 | 44.00 | 43.00 | ❌ | 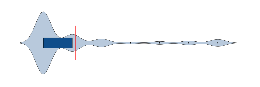 |
| N = 103 including n = 9 recompleted responses. BC-QoL= bronchiectasis child-specific parent-proxy quality of life, higher reversed BC-QoL scores indicate greater severity *^*^* during the past week, how worried/ concerned were you: *^†^* during the past week, how often:, ^‡^Scores reversed for impact analysis, means and medians are of reversed scores. | | | | | | | | | |

eTable 2: Item-total spearman correlations of item pairs with high (>0.75) item-item correlations.

| Item 1 | Item 2 | Correlation | Item 1 ITC | Item 2 ITC |
| --- | --- | --- | --- | --- |
| Worry | sorry | 0.84 | 0.80 | 0.78 |
| Normallife | effects | 0.84 | 0.77 | 0.89 |
| Anxious | overprotective | 0.83 | 0.79 | 0.84 |
| Upset | sorry | 0.82 | 0.72 | 0.78 |
| worry | anxious | 0.80 | 0.80 | 0.79 |
| sorry | overprotective | 0.79 | 0.78 | 0.84 |
| unwell | healthy | 0.78 | 0.82 | 0.83 |
| childtired | healthy | 0.78 | 0.83 | 0.83 |
| effects | lungdamage | 0.78 | 0.89 | 0.82 |
| overprotective | effects | 0.77 | 0.84 | 0.89 |
| unwell | worse | 0.76 | 0.82 | 0.75 |
| unwell | activities | 0.76 | 0.82 | 0.73 |
| worry | overprotective | 0.76 | 0.80 | 0.84 |
| effects | healthy | 0.76 | 0.89 | 0.83 |
| worry | upset | 0.76 | 0.80 | 0.72 |
| anxious | upset | 0.75 | 0.79 | 0.72 |
| anxious | sorry | 0.75 | 0.79 | 0.78 |
| sleep | childscough | 0.75 | 0.80 | 0.75 |

^#^ Item with lower item-total spearman correlation is highlighted in red

eTable 3: Factor analysis results: Item loading of oblimin rotated solution with three factors, items. Cross loading items highlighted red. Absolute loading value <0.3 denoted by great font colour.

|  | Factor_1 | Factor_2 | Factor_3 | Communality | Uniqueness | Complexity |
| --- | --- | --- | --- | --- | --- | --- |
| healthy | 1.001 | -0.100 | -0.066 | 0.87 | 0.13 | 1.03 |
| childtired | 0.894 | -0.018 | 0.056 | 0.78 | 0.22 | 1.01 |
| antibiotics | 0.875 | -0.118 | 0.019 | 0.63 | 0.37 | 1.04 |
| sleep | 0.858 | 0.018 | 0.297 | 0.86 | 0.14 | 1.24 |
| unwell | 0.831 | 0.086 | 0.060 | 0.81 | 0.19 | 1.03 |
| growth | 0.807 | -0.025 | 0.144 | 0.65 | 0.35 | 1.07 |
| appetite | 0.769 | -0.030 | 0.140 | 0.59 | 0.41 | 1.07 |
| childscough | 0.752 | 0.066 | 0.225 | 0.70 | 0.30 | 1.19 |
| exposed | 0.751 | 0.029 | -0.180 | 0.62 | 0.38 | 1.12 |
| worse | 0.749 | 0.073 | -0.057 | 0.65 | 0.35 | 1.03 |
| lungdamage | 0.712 | 0.166 | -0.278 | 0.77 | 0.23 | 1.42 |
| sport | 0.674 | 0.102 | -0.095 | 0.57 | 0.43 | 1.09 |
| effects | 0.666 | 0.308 | -0.234 | 0.88 | 0.12 | 1.69 |
| activities | 0.659 | 0.180 | -0.104 | 0.64 | 0.36 | 1.20 |
| normallife | 0.588 | 0.282 | -0.334 | 0.76 | 0.24 | 2.08 |
| judged | 0.581 | 0.295 | -0.040 | 0.67 | 0.33 | 1.49 |
| upset | -0.103 | 0.999 | 0.096 | 0.87 | 0.13 | 1.04 |
| sorry | 0.015 | 0.927 | 0.017 | 0.88 | 0.12 | 1.00 |
| worry | 0.003 | 0.889 | 0.014 | 0.79 | 0.21 | 1.00 |
| anxious | 0.083 | 0.827 | -0.029 | 0.79 | 0.21 | 1.02 |
| overprotective | 0.225 | 0.731 | -0.117 | 0.83 | 0.17 | 1.24 |
| doctors | 0.140 | 0.569 | -0.051 | 0.46 | 0.54 | 1.14 |
| awakened | 0.284 | 0.435 | 0.608 | 0.83 | 0.17 | 2.29 |

eTable 4: Eigenvalues, Variance Explained, and Factor Correlations for Rotated Factor Solution

| Property | Factor_1 | Factor_2 | Factor_3 |
| --- | --- | --- | --- |
| SS loadings | 10.442 | 5.581 | 0.886 |
| Proportion Variance | 0.454 | 0.243 | 0.039 |
| Cumulative Variance | 0.454 | 0.697 | 0.735 |
| Proportion Explained | 0.618 | 0.330 | 0.052 |
| Cumulative Proportion | 0.618 | 0.948 | 1.000 |
| Factor_1 | 1.000 | 0.719 | 0.030 |
| Factor_2 | 0.719 | 1.000 | 0.006 |
| Factor_3 | 0.030 | 0.006 | 1.000 |

eTable 5: Minimal important difference calculated using distribution methods. n = 78

|  | **Time-point 1** (Day 0) | | | | | **Time-point 2** (Day 21) | | | | |
| --- | --- | --- | --- | --- | --- | --- | --- | --- | --- | --- |
|  | Emotional | Physical | Social | Total | VCD | Emotional | Physical | Social | Total | VCD |
| Mean | 4.29 | 4.36 | 4.47 | 4.37 | 1.43 | 4.97 | 4.95 | 4.97 | 4.96 | 1.31 |
| SD^*^ | 1.57 | 1.70 | 1.49 | 1.49 | 1.50 | 1.72 | 1.70 | 1.58 | 1.64 | 1.28 |
| Cronbach's α | 0.93 | 0.93 | 0.90 | 0.97 | --- | --- | --- | --- | --- | --- |
| Minimal important difference calculations | | | | | | | | | | |
| Effect size | 0.43 | 0.35 | 0.34 | 0.40 | --- | --- | --- | --- | --- | --- |
| SEM^†^ | 0.42 | 0.45 | 0.47 | 0.26 | --- | --- | --- | --- | --- | --- |
| One-half SD^*^ | 0.79 | 0.85 | 0.74 | **0.74** | --- | --- | --- | --- | --- | --- |
| One-third SD^*^ | 0.52 | 0.57 | 0.50 | 0.50 | --- | --- | --- | --- | --- | --- |
| ^*^SD: Standard deviation ^†^SEM: Standard error of measurement | | | | | | | | | | |

eTable 6: Minimal important difference calculated using anchor-based methods. Changes in verbal category descriptive (VCD) cough score used as anchor. n= 65 participants with BC-QoL and cough responses at day 0 and day 21.

|  | Change in VCD score between day 0 and day 21 | | | | |
| --- | --- | --- | --- | --- | --- |
|  | No change 0 (n = 23) | Small change  ±1 (n = 22) | Moderate change  ±2 (n = 9) | Large change  ±3 (n = 7) | Extreme change  ±4 (n = 4) |
| Emotional | 1.01 (1) | 1.32 (1.27) | 0.92 (0.8) | 1.23 (0.92) | 0.75 (0.4) |
| Physical | 0.85 (0.86) | 1.44 (1.23) | 1.06 (1.05) | 1.68 (1.15) | 0.69 (0.39) |
| Social | 0.74 (0.72) | 1.29 (0.96) | 0.95 (0.67) | 1.1 (0.84) | 0.57 (0.4) |
| Total | 0.82 (0.75) | 1.32 (1.02) | 0.77 (0.82) | 1.31 (0.77) | 0.67 (0.3) |
| N = 65 participants with both BE-QoL and cough score at enrolment and follow-up Change in BE-QoL scores between time-point 1 & 2, mean(SD). | | | | | |

### References

1.	Anderson-James S, Newcombe PA, Marchant JM, O'grady KA, Acworth JP, Stone DG*, et al.* An acute cough-specific quality-of-life questionnaire for children: Development and validation. *J Allergy Clin Immunol*. 2015;135(5):1179-85 e1-4. doi:10.1016/j.jaci.2014.08.036.

2.	Newcombe PA, Sheffield JK, Juniper EF, Marchant JM, Halsted RA, Masters IB*, et al.* Development of a parent-proxy quality-of-life chronic cough-specific questionnaire: clinical impact vs psychometric evaluations. *Chest*. 2008;133(2):386-95. doi:10.1378/chest.07-0888.

3.	Marchant JM, Cook AL, Roberts J, Yerkovich ST, Goyal V, Arnold D*, et al.* Burden of Care for Children with Bronchiectasis from Parents/Carers Perspective. *J Clin Med*. 2021;10(24):5856. doi:10.3390/jcm10245856.

4.	Newcombe PA, Sheffield JK, Chang AB. Minimally important change in a Parent-Proxy Quality-of-Life questionnaire for pediatric chronic cough. *Chest*. 2011;139(3):576-80. doi:10.1378/chest.10-1476.

5.	Newcombe PA, Sheffield JK, Juniper EF, Petsky HL, Willis C, Chang AB. Validation of a parent-proxy quality of life questionnaire for paediatric chronic cough (PC-QOL). *Thorax*. 2010;65(9):819-23. doi:10.1136/thx.2009.133868.

6.	Newcombe PA, Sheffield JK, Petsky HL, Marchant JM, Willis C, Chang AB. A child chronic cough-specific quality of life measure: development and validation. *Thorax*. 2016;71(8):695-700. doi:10.1136/thoraxjnl-2015-207473.

7.	Quittner AL, Marciel KK, Salathe MA, O'donnell AE, Gotfried MH, Ilowite JS*, et al.* A preliminary quality of life questionnaire-bronchiectasis: a patient-reported outcome measure for bronchiectasis. *Chest*. 2014;146(2):437-48. doi:10.1378/chest.13-1891.

8.	Quittner AL, O'donnell AE, Salathe MA, Lewis SA, Li X, Montgomery AB*, et al.* Quality of Life Questionnaire-Bronchiectasis: final psychometric analyses and determination of minimal important difference scores. *Thorax*. 2015;70(1):12-20. doi:10.1136/thoraxjnl-2014-205918.

9.	Mills DR, Masters IB, Yerkovich ST, Mceniery J, Kapur N, Chang AB*, et al.* Radiographic Outcomes in Pediatric Bronchiectasis and Factors Associated with Reversibility. *Am J Respir Crit Care Med*. 2024;210(1):97-107. doi:10.1164/rccm.202402-0411OC.

10.	Harris PA, Taylor R, Thielke R, Payne J, Gonzalez N, Conde JG. Research electronic data capture (REDCap)--a metadata-driven methodology and workflow process for providing translational research informatics support. *J Biomed Inform*. 2009;42(2):377-81. doi:10.1016/j.jbi.2008.08.010.

11.	Chang AB, Newman RG, Carlin JB, Phelan PD, Robertson CF. Subjective scoring of cough in children: parent-completed vs child-completed diary cards vs an objective method. *Eur Respir J*. 1998;11(2):462-6. doi:10.1183/09031936.98.11020462.

12.	Hays RD, Morales LS. The RAND-36 measure of health-related quality of life. *Ann Med*. 2001;33(5):350-7. doi:10.3109/07853890109002089.

13.	Kapur N, Masters IB, Newcombe P, Chang AB. The burden of disease in pediatric non-cystic fibrosis bronchiectasis. *Chest*. 2012;141(4):1018-24. doi:10.1378/chest.11-0679.

14.	Henry JD, Crawford JR. The short-form version of the Depression Anxiety Stress Scales (DASS-21): construct validity and normative data in a large non-clinical sample. *Br J Clin Psychol*. 2005;44(Pt 2):227-39. doi:10.1348/014466505X29657.

15.	Varni JW, Burwinkle TM, Seid M, Skarr D. The PedsQL 4.0 as a pediatric population health measure: feasibility, reliability, and validity. *Ambul Pediatr*. 2003;3(6):329-41. doi:10.1367/1539-4409(2003)003<0329:tpaapp>2.0.co;2.

16.	Juniper EF, Guyatt GH, Streiner DL, King DR. Clinical impact versus factor analysis for quality of life questionnaire construction. *J Clin Epidemiol*. 1997;50(3):233-8. doi:10.1016/s0895-4356(96)00377-0.

17.	Kaiser HF. An index of factorial simplicity. *Psychometrika*. 1974;39(1):31-6.

18.	Watkins MW. Exploratory Factor Analysis: A Guide to Best Practice. *Journal of Black Psychology*. 2018;44(3):219-46. doi:10.1177/0095798418771807.

19.	Bartlett MS. A note on the multiplying factors for various χ 2 approximations. *Journal of the Royal Statistical Society Series B (Methodological)*. 1954;16(2):296-8. <Go to ISI>://WOS:A1954YH07100016

20.	Cattell RB. The Scree Test For The Number Of Factors. *Multivariate Behav Res*. 1966;1(2):245-76. doi:10.1207/s15327906mbr0102_10.

21.	Cronbach LJ. Coefficient alpha and the internal structure of tests. *Psychometrika*. 1951;16(3):297-334. <Go to ISI>://WOS:000203834000004

22.	Spearman C. Correlation Calculated from Faulty Data. *British Journal of Psychology, 1904-1920*. 2011;3(3):271-95. doi:10.1111/j.2044-8295.1910.tb00206.x.

23.	Yost KJ, Eton DT. Combining distribution- and anchor-based approaches to determine minimally important differences: the FACIT experience. *Eval Health Prof*. 2005;28(2):172-91. doi:10.1177/0163278705275340.

24.	Mundfrom DJ, Shaw DG, Ke TL. Minimum sample size recommendations for conducting factor analyses. *International journal of testing*. 2005;5(2):159-68.

25.	R Core Team. R: A language and environment for statistical computing. Vienna, Austria: R Foundation for Statistical Computing; 2023.

26.	Sjoberg DD, Whiting K, Curry M, Lavery JA, Larmarange J. Reproducible Summary Tables with the gtsummary Package. *The R Journal*. 2021;13(1):570. doi:10.32614/rj-2021-053.

27.	Revelle W, Revelle MW. Package ‘psych’. *The comprehensive R archive network*. 2015;337(338):161-5.

28.	Caldwell AR. SimplyAgree: an R package and jamovi module for simplifying agreement and reliability analyses. *Journal of Open Source Software*. 2022;7(71):4148.
