## Supplementary material for "Development and validation of a parent proxy bronchiectasis child quality of life instrument: The BC-QoL": eAppendix-2

**BC-QOL – Parent Proxy***

The following items relate to some of the ways in which parents might respond towards their child’s bronchiectasis. Please consider each and respond by marking the box that best corresponds to your own thoughts, feelings, and behaviours towards your child’s bronchiectasis.

| ***During the last week,***  ***how often:*** | **All the time** | **Most of the time** | **Quite often** | **Some of the time** | **Once in a while** | **Hardly any of the time** | **None of the time** |
| --- | --- | --- | --- | --- | --- | --- | --- |
| 1. Did you feel upset because of your child’s bronchiectasis? | □ | □ | □ | □ | □ | □ | □ |
| 2. Were you awakened during the night because of your child’s cough from his/her bronchiectasis? | □ | □ | □ | □ | □ | □ | □ |
| 3. Did you feel sorry for your child because of his/her bronchiectasis? | □ | □ | □ | □ | □ | □ | □ |
| 4. Did you feel overly protective toward your child because of his/her bronchiectasis? | □ | □ | □ | □ | □ | □ | □ |
| 5. Did you feel anxious because of his/her bronchiectasis? | □ | □ | □ | □ | □ | □ | □ |
| 6. Did you worry about your child’s bronchiectasis? | □ | □ | □ | □ | □ | □ | □ |
| 7. Were you concerned that general doctors did not know how to treat your child’s bronchiectasis? | □ | □ | □ | □ | □ | □ | □ |

| ***During the last week, how worried or concerned were you:*** | **Very very** | **Very** | **Fairly** | **Some-what** | **A little** | **Hardly** | **Not** |
| --- | --- | --- | --- | --- | --- | --- | --- |
|  | **Worried/ concerned** | | | | | | |
| 8. About your child’s bronchiectasis becoming worse/progressing? | □ | □ | □ | □ | □ | □ | □ |
| 9. About your child being able to do normal activities such as playing and school because of his/her bronchiectasis? | □ | □ | □ | □ | □ | □ | □ |
| 10. About your child feeling unwell because of his/her bronchiectasis? | □ | □ | □ | □ | □ | □ | □ |
| 11. About your child’s bronchiectasis symptoms bothering or being judged by other people? | □ | □ | □ | □ | □ | □ | □ |
| 12. About your child’s ability to lead a normal life as an adult? | □ | □ | □ | □ | □ | □ | □ |
| 13. About the effects of your child’s bronchiectasis on him/her? | □ | □ | □ | □ | □ | □ | □ |
| 14. About your child’s cough? | □ | □ | □ | □ | □ | □ | □ |
| 15. About your child not sleeping well because of the bronchiectasis? | □ | □ | □ | □ | □ | □ | □ |
| 16. About your child being exposed to others who may be ill? | □ | □ | □ | □ | □ | □ | □ |
| 17. About your child not being able to play sport and exercise like other children? | □ | □ | □ | □ | □ | □ | □ |
| 18. About your child feeling healthy? | □ | □ | □ | □ | □ | □ | □ |
| 19. About your child’s growth and weight? | □ | □ | □ | □ | □ | □ | □ |
| 20. About your child feeling tired because of their bronchiectasis? | □ | □ | □ | □ | □ | □ | □ |
| 21. About your child taking too many antibiotics? | □ | □ | □ | □ | □ | □ | □ |
| 22. About long term damage to your child’s lungs because of the bronchiectasis? | □ | □ | □ | □ | □ | □ | □ |
| 23. About your child’s appetite? | □ | □ | □ | □ | □ | □ | □ |
| **Is there anything further you would like to comment on in relation to your child’s bronchiectasis and its impact on you and your child or family’s lives?**  **………………………………………………………………………………………………………………………………………**  **……………………………………………………………………………………………………………………………………….** | | | | | | | |

**Thank you for your participation**

***- Not to be amended or changed in any manner without express consent of authors -**
